# Lysophosphatidylcholines are associated with P-tau181 levels in early stages of Alzheimer’s Disease

**DOI:** 10.1101/2023.08.24.23294581

**Authors:** Vrinda Kalia, Dolly Reyes-Dumeyer, Saurabh Dubey, Renu Nandakumar, Annie J. Lee, Rafael Lantigua, Martin Medrano, Diones Rivera, Lawrence S. Honig, Richard Mayeux, Gary W. Miller, Badri N. Vardarajan

**Affiliations:** Department of Environmental Health Sciences, Mailman School of Public Health, Columbia University. 722 West 168th Street, New York, NY 10032; Taub Institute for Research on Alzheimer’s Disease and the Aging Brain, College of Physicians and Surgeons, Columbia University. 630 West 168th Street, New York, NY 10032; The Gertrude H. Sergievsky Center, College of Physicians and Surgeons, Columbia University. 630 West 168th Street, New York, NY 10032; Department of Medicine, College of Physicians and Surgeons, Columbia University, and the New York Presbyterian Hospital. 630 West 168th Street, New York, NY 10032; School of Medicine, Pontificia Universidad Católica Madre y Maestra, Santiago, Dominican Republic; Department of Neurosurgery, CEDIMAT, Plaza de la Salud, Santo Domingo, Dominican Republic.; Department of Neurology, College of Physicians and Surgeons, Columbia University and the New York Presbyterian Hospital. 710 West 168th Street, New York, NY 10032; Department of Epidemiology, Mailman School of Public Health, Columbia University. 722 West 168th Street, New York, NY 10032

**Author notes:** To whom correspondence should be addressed, Address Correspondence to: Badri N. Vardarajan, PhD Taub Institute, Columbia University, 620 West 168th Street New York, NY 10032, Gary Miller, PhD Dept of Environmental Health Sciences, Columbia University, 722 West 168th Street, New York, NY 10032.

## Abstract

**Background:** We investigated systemic biochemical changes in Alzheimer’s disease (AD) by investigating the relationship between circulating plasma metabolites and both clinical and biomarker-assisted diagnosis of AD.

**Methods:** We used an untargeted approach with liquid chromatography coupled to high-resolution mass spectrometry to measure exogenous and endogenous small molecule metabolites in plasma from 150 individuals clinically diagnosed with AD and 567 age-matched elderly without dementia of Caribbean Hispanic ancestry. Plasma biomarkers of AD were also measured including P-tau181, Aβ40, Aβ42, total tau, neurofilament light chain (NfL) and glial fibrillary acidic protein (GFAP). Association of individual and co-expressed modules of metabolites were tested with the clinical diagnosis of AD, as well as biologically-defined AD pathological process based on P-tau181 and other biomarker levels.

**Results:** Over 4000 metabolomic features were measured with high accuracy. First principal component (PC) of lysophosphatidylcholines (lysoPC) that bind to or interact with docosahexaenoic acid (DHA), eicosapentaenoic acid (EPA) and arachidonic acid (AHA) was associated with decreased risk of AD (OR=0.91 [0.89-0.96], p=2e-04). Restricted to individuals without an *APOE* ε*4 allele* (OR=0.89 [0.84-0.94], p= 8.7e-05), the association remained. Among individuals carrying at least one *APOE* ε*4* allele, PC4 of lysoPCs moderately increased risk of AD (OR=1.37 [1.16-1.6], p=1e-04). Essential amino acids including tyrosine metabolism pathways were enriched among metabolites associated with P-tau181 levels and heparan and keratan sulfate degradation pathways were associated with Aβ42/Aβ40 ratio reflecting different pathways enriched in early and middle stages of disease.

**Conclusions:** Our findings indicate that unbiased metabolic profiling can identify critical metabolites and pathways associated with β-amyloid and phosphotau pathology. We also observed an *APOE* ε*4* dependent association of lysoPCs with AD and that biologically-based diagnostic criteria may aid in the identification of unique pathogenic mechanisms.

**Graphical Abstract:** 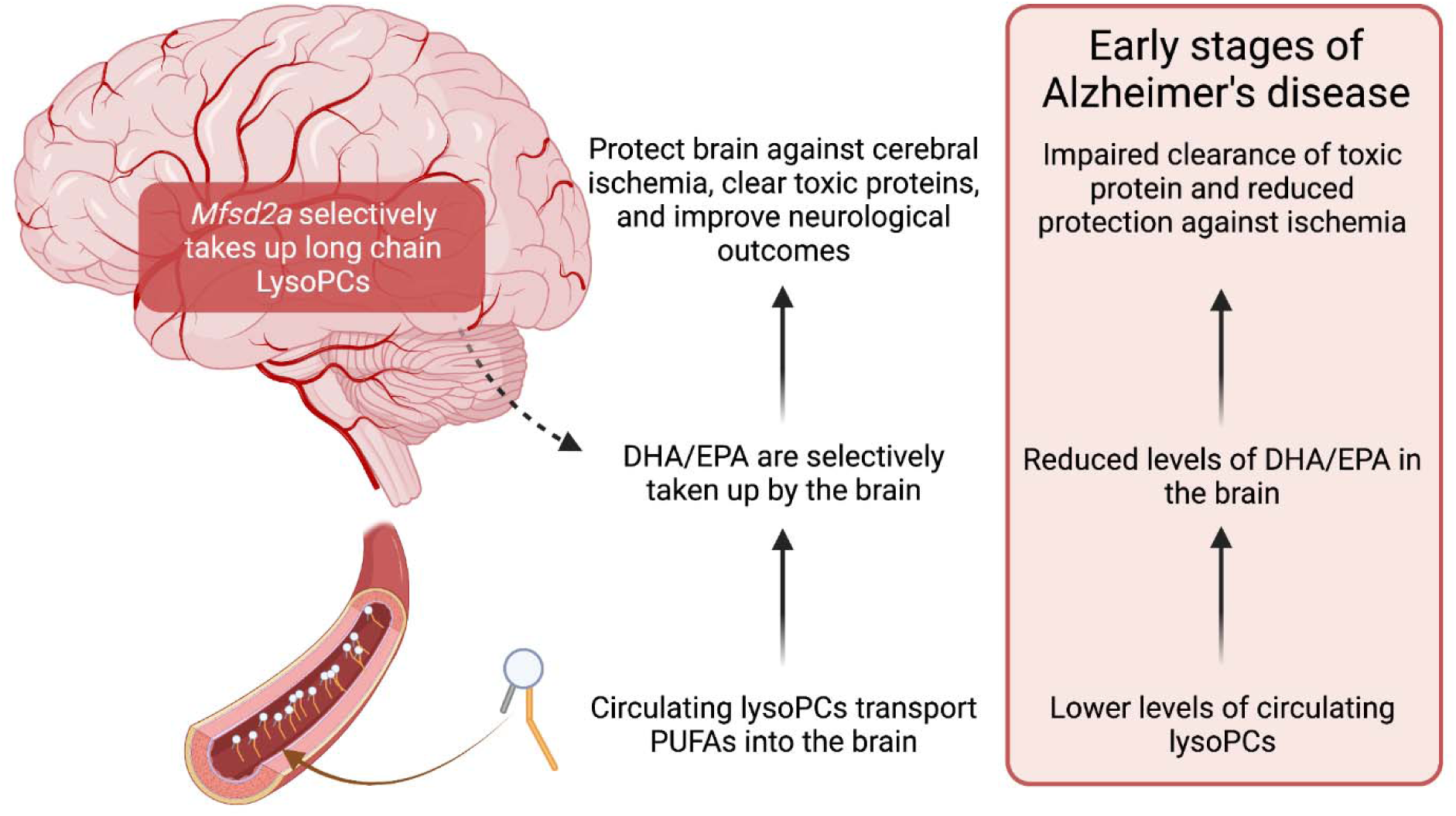

## Introduction

Alzheimer’s disease (AD) is a progressive neurodegenerative disorder characterized by cognitive and memory decline, affecting millions of individuals worldwide. Despite extensive research, the underlying pathogenic mechanisms of AD have not been fully revealed, hindering the development of effective therapeutic strategies. However, recent advancements in high-throughput omics technologies have provided a powerful platform to explore the complex molecular landscape of AD^1^.

Mass spectrometry-based metabolic profiling, a.k.a. metabolomics, offers a comprehensive analysis of small molecules involved in cellular metabolism. It provides a unique opportunity to unravel metabolic alterations associated with disease pathogenesis, thus contributing to a better understanding of AD at the molecular level^2–5^. Untargeted metabolomics allows the unbiased profiling of the entire metabolome, including both known and unknown metabolites.

Metabolomic studies in AD have revealed a range of altered metabolic signaling. Several studies have demonstrated dysregulation of energy metabolism pathways in AD^6–8^. These alterations involve changes in glucose metabolism^9^, including reduced glycolysis^10–14^ and impaired mitochondrial function^15–17^, and decreased levels of metabolites such as glucose, lactate, and pyruvate^18^. Alterations in the tricarboxylic acid (TCA) cycle intermediates, have also been observed. Studies have reported lower levels of phosphatidylcholine, phosphatidylethanolamine, and sphingomyelins in AD^19–22^, suggesting disruptions in membrane integrity and signaling pathways, altered cholesterol metabolism has also been implicated. Other studies have uncovered alterations in amino acid metabolism in AD^23^. Reduced levels of certain amino acids, such as tryptophan^24^, phenylalanine^25,26^, tyrosine^27,28^, and branched-chain amino acids (valine, leucine, isoleucine)^3,29–38^ may reflect disruptions in neurotransmitter synthesis, neuroinflammation, and protein homeostasis. Studies have also shown alterations in the levels of neurotransmitters such as acetylcholine, glutamate, and γ-aminobutyric acid (GABA) in AD patients^39–43^. These changes may contribute to cognitive dysfunction and synaptic alterations in the disease.

Several groups have reported elevated levels of reactive oxygen species (ROS) and oxidative damage markers, along with alterations in antioxidant metabolites and enzymes, have been observed in whole blood and brains of AD patients. These findings suggest a role for altered redox status in AD pathogenesis^44–47^.

Given that metabolomics is the omics layer closest to the phenotype, it has the potential to uncover critical insights into the disease risk and progression, and potentially uncover therapeutic targets. By integrating metabolomics data with clinical diagnosis and plasma biomarker levels of AD, we aim to identify metabolic networks underlying the disease. In this study, we investigated the association between metabolites and clinical and biomarker assisted diagnosis of AD to detect early and mid-stage metabolic changes in disease.

## Methods

### Participants

The Estudio Familiar de Influencia Genetica en Alzheimer (EFIGA) has been recruiting individuals with suspected sporadic and familial AD and healthy controls similar in age through advertisements in local newspapers and radio stations, and through clinical referrals in the Dominican Republic and in the Washington Heights neighborhood of New York City^48^. Participants in this study provided informed consent under protocols approved by the Columbia University Irving Medical Center Institutional Review Board, and the National Health Bioethics

Committee of the Dominican Republic (CONABIOS). They underwent a medical and neurological interview for history and detailed examinations, a neuropsychological test battery, and collection of blood for plasma and DNA processing. Brain tomography or magnetic resonance as well as CSF were performed in a subgroup of participants. The diagnosis of Alzheimer’s clinical syndrome according to NIA-AA criteria^49^. All clinical diagnosis were determined in a consensus conference attended by a neurologist, a neuropsychologist, and an internist with expertise in dementia and geriatrics. For the analyses in this manuscript, biological samples and data from individuals recruited between January 1, 2018, and April 30, 2022, were considered.

### Sample collection

Blood was collected in K2EDTA tubes by standard venipuncture and transported to a laboratory for centrifugation, preparation of plasma, and storage at −80°C within 2 hours of collection. CSF was obtained by standard aseptic technique, distributed into aliquots of 400 µL each in polypropylene tubes, frozen, and stored at −80°C^48^.

### Plasma and CSF metabolomics data generation

Plasma and CSF metabolites were extracted using acetonitrile and the extracts were injected in triplicate on two chromatographic columns: a hydrophilic interaction column (HILIC) under positive ionization (HILIC+)^50^ and a C18 column under negative ionization (C18-)^51^ coupled to a Thermo Orbitrap HFX Q-Exactive mass spectrometer, scanning for molecules within 85 – 1250 kDa. This produced three technical replicates per sample per column. The untargeted mass spectral data were processed through a computational pipeline that leverages open source feature detection and peak alignment software, apLCMS^52^ and xMSanalyzer^53^. The feature tables were generated containing information on the mass-to-charge (m/z) ratio, retention time, and median summarized abundance/intensity of each ion for each sample. Correction for batch effects was performed using ComBat, which uses an empirical Bayesian framework to adjust for known batches in which the samples were run^54^. Each of these ions are referred to as metabolic features. For the analysis, metabolic features detected in at least 70% of the samples were retained, leaving 3253 features from the HILIC+ column and 3628 features from the C18-column for plasma samples and 4460 features from the HILIC+ column and 4501 features from the C18-column for CSF samples. Zero-intensity values were considered below the detection limit of the instrument and were imputed with half the minimum intensity observed for each metabolic feature. The intensity of each metabolic feature was log10 transformed, quantile normalized, and auto-scaled for normalization and standardization.

### Metabolite annotation

Annotations were made using an internal library and by matching to the Human Metabolome Database (HMDB) using the R package xMSannotator (version 1.3.2)^53^. This uses a multistage clustering algorithm method to determine metabolic pathway associations, intensity profiles, retention time, mass defect, and isotope/adduct patterns to assign putative annotations to metabolic features. When a feature had multiple matches, we used the following rules to assign an annotation: first, we screened features based on the confidence score assigned by xMSannotator, and the annotation with the highest score was used. Second, if all annotations had the same score, we chose the annotation with the lowest difference in expected and observed mass (delta parts per million (ppm)). Finally, if all features had the same score and delta ppm, we indicated the identity as “multiple matches” since we couldn’t decipher a unique putative annotation. If a feature did not match any database entries, it was denoted as “unknown” (33% from HILIC + column and 40% from C18 – column). The confidence in annotation was based on criteria defined by Schymanski et al^55^, where level 1 corresponds to a confirmed structure identified through MS/MS and/or comparison to an authentic standard; level 2 to a probable structure identified through spectral matches to a database; level 3 to a putative identification with a speculative structure; level 4 to an unequivocal molecular formula but with insufficient evidence to propose a structure; and level 5 to an exact mass but not enough information to assign a formula.

### Blood based biomarker analyses

The methods have been previously described in detail^48^. Briefly, the plasma biomarkers assays were performed in duplicate using the SIMOA HD-X platform. Neurology 3-Plex A kits were used to determine levels of Aβ42, Aβ40, and T-tau, the Advantage V2 kit for P-tau181, and the Neurology 2-Plex B for GFAP and NfL. Ratio of Aβ42/Aβ40 was also calculated.

### Biomarker positive for AD

Based on previous analysis^48^, P-tau-181 plasma level < 2.33 considered biomarker status negative and ≥ 2.33 considered biomarker status positive. We use biomarker positive and biological AD interchangeably in the manuscript.

### Statistical analysis approach

We used two approaches to find circulating metabolic features associated with outcomes of interest: 1) a metabolome-wide association study (MWAS) framework with correction for multiple comparisons by controlling the false discovery rate (FDR) at 5%, and 2) a co-expression analysis to find modules of metabolic features associated with outcomes, providing a means of unsupervised dimensionality reduction based on correlation between the metabolic features. Both analyses were conducted separately for data from each column. All analyses were conducted in R (version 3.6.3).

### Metabolome-wide association study (MWAS)

MWAS was conducted using multiple linear models, adjusted for age and sex. The analyses were conducted separately for data from each column. We corrected for multiple comparisons using an FDR of 5% and q-values were estimated using the Benjamini-Hochberg (BH) method.

### Metabolite Co-abundance analysis

Co-abundance modular analysis was conducted using weighted gene correlation network analysis^56^ using the WGCNA R package (version 1.69). Using normalized intensity values for each metabolic feature from each sample, we first constructed a metabolic feature co-expression network using pairwise Pearson correlations between each metabolic feature. We used a soft threshold of 4 for the HILIC+ data and 3 for the C18 – data, chosen based on saturation of the R^2^ at 0.9. This correlation network, where the nodes are metabolic features and edges are the scaled correlation coefficients, was used to create the topological overlap matrix (TOM), which provides a measure of similarity between a given pair of metabolic features in the network. This similarity matrix was used to create a dendrogram to assign metabolic features into modules based on their co-expression pattern. We used the following parameters: minimum module size of 30, merge cutHeight of 0.25, an unsigned network, and a reassign threshold of 0. After network and dendrogram construction, modules were defined using the *moduleEigengenes* function in WGCNA. The module eigengene is a quantitative representation of a module derived from a principal component analysis (PCA) as the first PC, conducted using only those metabolic features that were part of the module. Association analyses were conducted to find modules associated with outcomes in linear regression models, adjusted for age and sex. We used the Bonferroni method to correct for multiple comparisons.

### Subgroup comparisons

Subgroup comparisons were conducted using logistic regression and multinomial logistic regression using the R package nnet (version 7.3-12). We created three different models to compare the six different subgroups: a) **BM+/Cases, BM+/Controls and BM-/cases vs BM-/Controls-**using healthy participants with biomarker status negative (BM-) as the reference (i.e BM-/controls), we compared metabolic features with differential levels in participants with i) biomarker status positive and a clinical diagnosis of AD (BM+/Cases), ii) biomarker status negative and a clinical diagnosis of AD (BM-/Cases) and biomarker positive with no clinical diagnosis of AD (BM+/Controls); b) **BM+/Case and BM+/Control vs BM-/Control**-metabolic features with differential levels among BM+/control, BM+/case, compared to BM-/control; and c) **BM+/case compared to BM+/control**.

### Pathway analysis

To determine the biological relevance of the metabolic features associated with AD and biomarkers, we conducted pathway analysis using the “functional analysis” module in MetaboAnalyst (version 5.0, ref)^57^, a web-based interface for comprehensive metabolomic data analysis. We used the MWAS results from both columns and applied a nominal p-value cut-off of 0.01 to determine metabolic pathway enrichment using the mummichog algorithm and the human MFN reference database^58^. We present results for pathways with a Fisher’s exact test p-value < 0.3.

### Chemical class enrichment

This approach was used to determine the different chemical classes represented by metabolic feature members of WGCNA modules significantly associated with outcomes. The main chemical classes enriched was determined using the Enrichment Analysis module in MetaboAnalyst using the HMDBIDs for features with an annotation confidence score < 3.

### Construction of lysophosphatidylcholine (lysoPCs) components and stratified analysis by ***APOE-****ε****4* status**

Based on findings described below we performed principal component analyses using all features that were annotated as lysoPCs from both columns (44 from HILIC + and 13 from C18-). Since PCs 1-5 explained ∼60% of the variance in the data, we used the first five PCs in logistic regression models to find the association with clinical diagnosis of AD and biomarker positive status, adjusted for age and sex. We tested for the presence of an interaction between the combinations of the lysoPCs and the presence of at least one *APOE-ε4* allele and performed a stratified analysis since there was a significant interaction term between *APOE-ε4* allele and PC4.

### Correlation between plasma and CSF metabolites

Among people with both plasma and CSF metabolomics data available (n = 113), plasma metabolites with level 1 confidence that were significantly associated with any outcome were tested for their correlation with the same metabolite identified in CSF using spearman correlation.

### Association between lysoPCs and brain pathology in the ROSMAP cohort

To provide external validation of our findings with lysoPCs, we obtained data from the ROSMAP cohort and examined associations between lysoPCs and brain pathology. We identified metabolites identified as LysoPCs and computed principal components as described above. Amongst the 42 PCs generated, we tested association of the top five with amyloid, tangles, total global pathology, clinical and pathological diagnosis of Alzheimer’s disease (AD).

## Results

### Study participants

717 participants were included in the study of whom 150 (20.9%) were diagnosed with clinical AD and 567 were cognitively unimpaired controls (Table 1). The study population had a mean age of 69.6 years (standard deviation (SD) = 7.6), the individuals with clinical AD were slightly older, with a mean age of 73.2 (SD = 8.3), compared to controls who had a mean age of 68.6 (SD = 7.2). Two-thirds of the group were women (65%) and this proportion was similar among AD patients (67%) and controls (65%). A third of the study group had at least one *APOE-ε4* allele (38%) and this proportion was only marginally higher in AD (43%) compared to controls (36%). Among AD, 58% were biomarker positive, while 29% of controls were biomarker positive. The mean levels of most plasma-based AD biomarkers were higher in AD than in controls, including P-tau181 (3.02 pg/mL (SD = 1.7) in AD and 2.13 pg/mL (SD = 1) in healthy controls), NfL pg/mL (26.4 (SD = 20.5) in AD and 17.3 pg/mL (SD = 19.2) in healthy controls), and GFAP pg/mL (219 (163) in AD and 140 pg/mL (96) in healthy controls). The mean ratio of αβ42/αβ40 was nominally lower in AD cases (0.049 (SD = 0.01)) compared to healthy controls (0.053 (SD = 0.03)). A subset of the study population, n = 113, also had CSF metabolomic data generated (S Table 1). Among them, 35 were clinically diagnosed with AD and 78 were controls. We also obtained postmortem brain tissue metabolomic data from a subset of participants from the ROSMAP cohort, n = 110 (S Table 2). Of them, 71 were diagnosed with AD and had brain pathology information available.

**Table 1.**
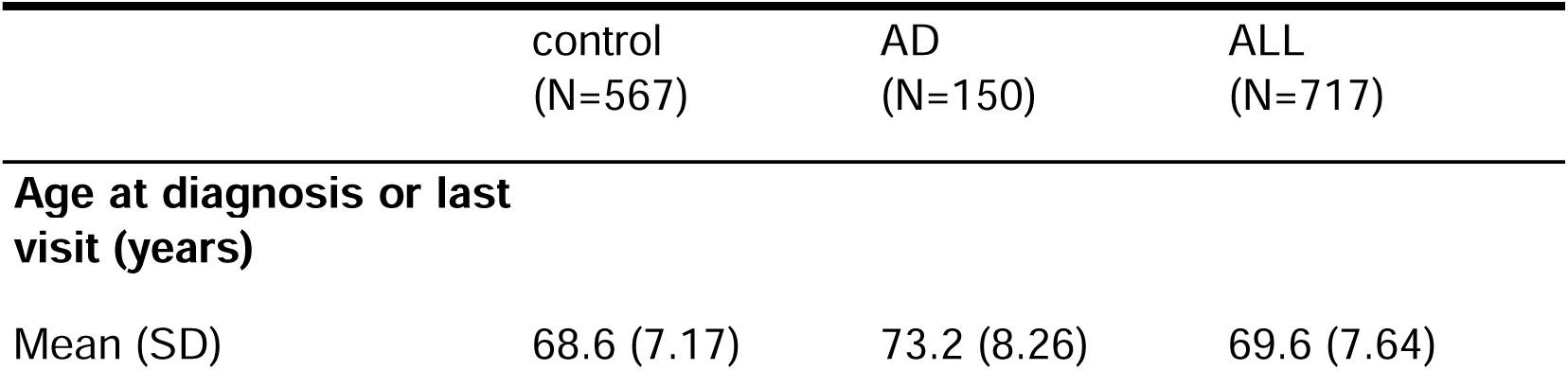
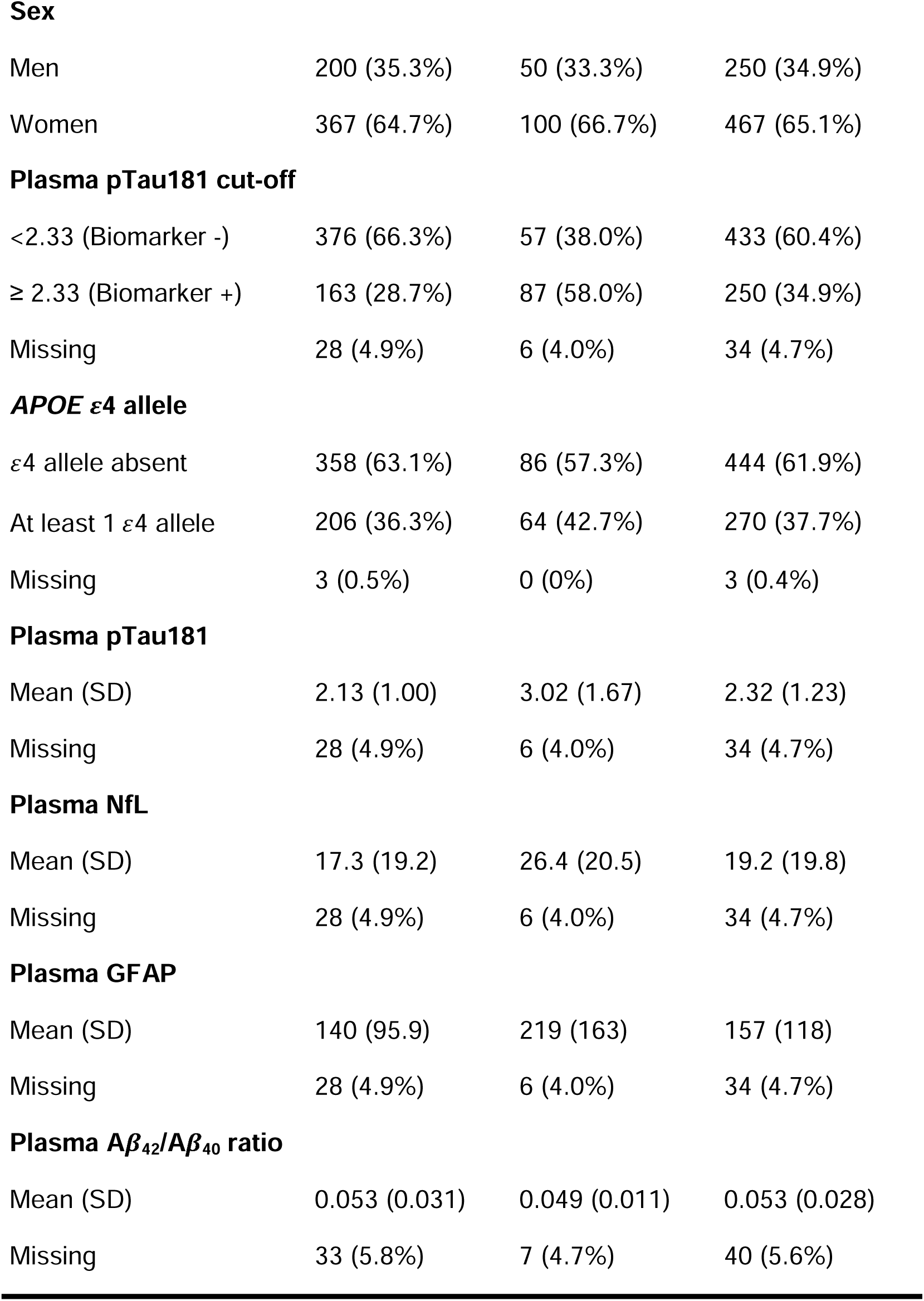
Characteristics of the study population.

|  | control<br>(N=567) | AD<br>(N=150) | ALL<br>(N=717) |
| --- | --- | --- | --- |
| **Age at diagnosis or last visit (years)** |  |  |  |
| Mean (SD) | 68.6 (7.17) | 73.2 (8.26) | 69.6 (7.64) |
| **Sex** |  |  |  |
| Men | 200 (35.3%) | 50 (33.3%) | 250 (34.9%) |
| Women | 367 (64.7%) | 100 (66.7%) | 467 (65.1%) |
| **Plasma pTau181 cut-off** |  |  |  |
| <2.33 (Biomarker -) | 376 (66.3%) | 57 (38.0%) | 433 (60.4%) |
| ≥ 2.33 (Biomarker +) | 163 (28.7%) | 87 (58.0%) | 250 (34.9%) |
| Missing | 28 (4.9%) | 6 (4.0%) | 34 (4.7%) |
| **APOE ε4 allele** |  |  |  |
| ε4 allele absent | 358 (63.1%) | 86 (57.3%) | 444 (61.9%) |
| At least 1 ε4 allele | 206 (36.3%) | 64 (42.7%) | 270 (37.7%) |
| Missing | 3 (0.5%) | 0 (0%) | 3 (0.4%) |
| **Plasma pTau181** |  |  |  |
| Mean (SD) | 2.13 (1.00) | 3.02 (1.67) | 2.32 (1.23) |
| Missing | 28 (4.9%) | 6 (4.0%) | 34 (4.7%) |
| **Plasma NfL** |  |  |  |
| Mean (SD) | 17.3 (19.2) | 26.4 (20.5) | 19.2 (19.8) |
| Missing | 28 (4.9%) | 6 (4.0%) | 34 (4.7%) |
| **Plasma GFAP** |  |  |  |
| Mean (SD) | 140 (95.9) | 219 (163) | 157 (118) |
| Missing | 28 (4.9%) | 6 (4.0%) | 34 (4.7%) |
| **Plasma Aβ<sub>42</sub>/Aβ<sub>40</sub> ratio** |  |  |  |
| Mean (SD) | 0.053 (0.031) | 0.049 (0.011) | 0.053 (0.028) |
| Missing | 33 (5.8%) | 7 (4.7%) | 40 (5.6%) |

**Table 2.**
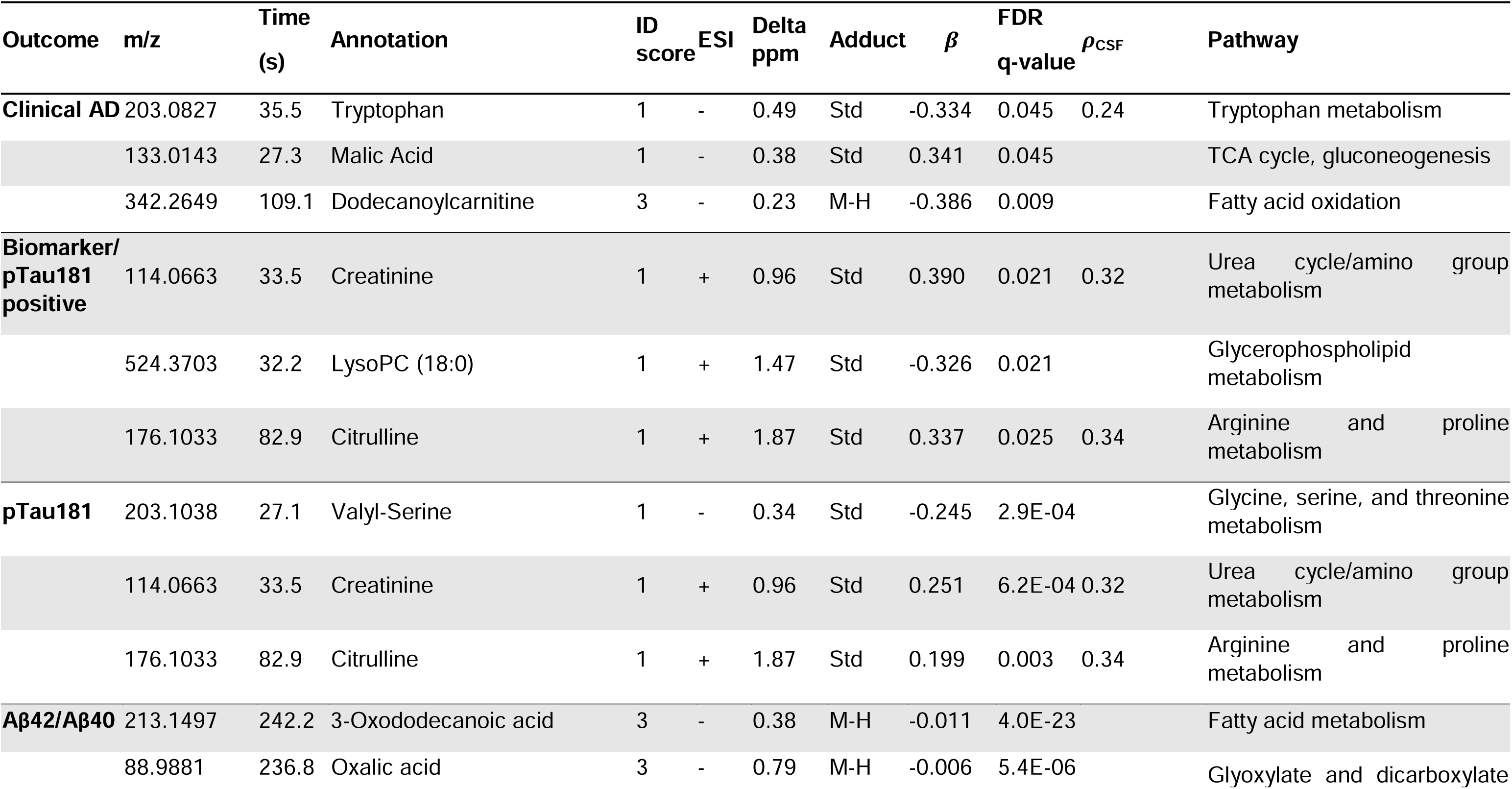
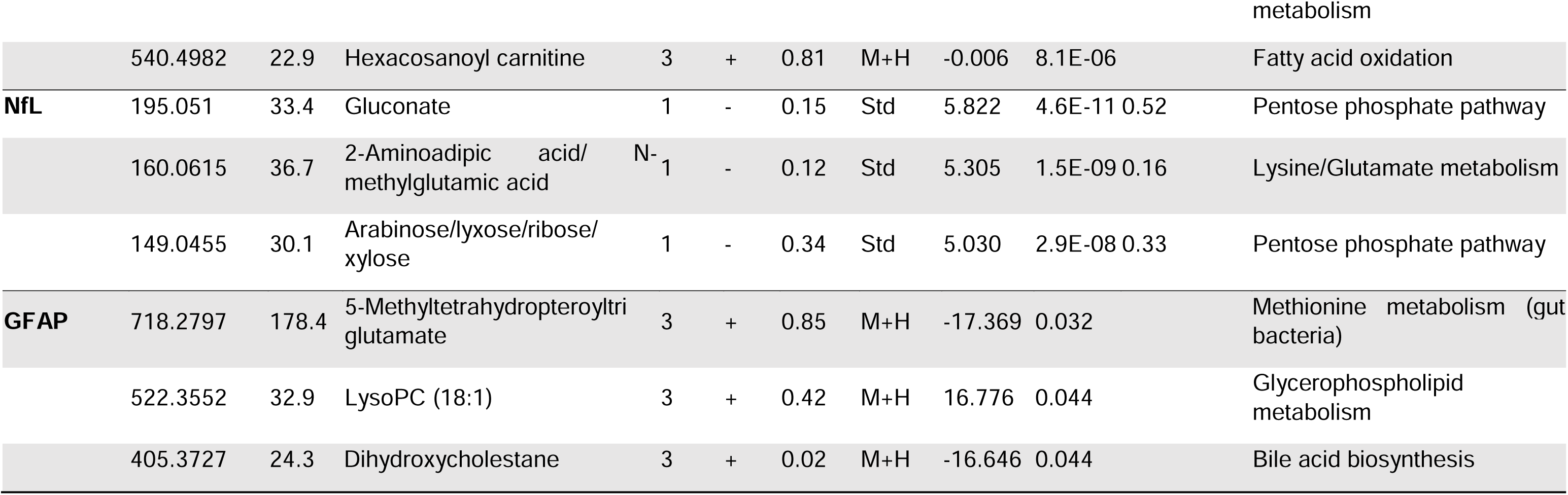
Metabolic features associated with outcomes investigated using a metabolome-wide association study framework. m/z: mass-to-charge ratio, Time: Retention time, ID score: confidence in annotation based on Schymanski scale (1 being the highest and 5 the lowest), ESI: electrospray ionization, Delta ppm: mass difference in parts per million, *ρ*_CSF_: correlation coefficient for metabolite measured in CSF (this was performed in a subset of participants, n =113). See supplemental table for complete list of features associated at FDR of 5%.

### Metabolome wide association study

We identified 6445 and 5827 metabolites in the HILIC+ and C18-columns respectively. Restricting to metabolites seen in at least 70% of the group, 3253 and 3628 metabolites were filtered for further analysis. 669 metabolites were associated with at least one phenotype (clinical diagnosis of AD, P-tau181 biomarker positive for AD or biological AD, plasma levels of αβ42/αβ 40 ratio, NfL, P-tau181 and GFAP). Of those, 174 metabolites were annotated with level 1 to level 3 confidence based on Schymanski scale (Figure 1, Table 2, supplementary Table 1).

**Figure 1.**
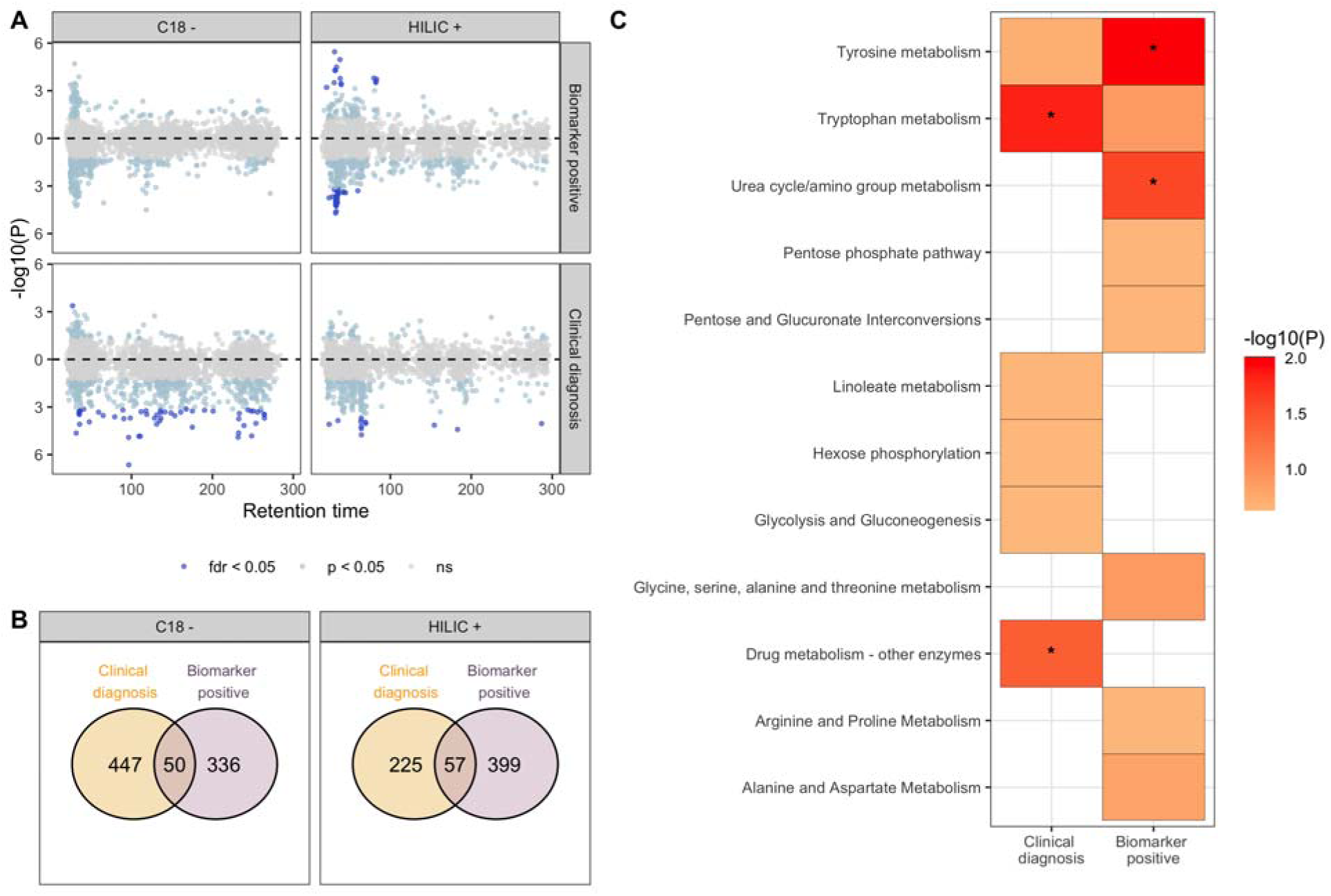
Metabolic features and pathways associated with clinical AD and with biomarker positive status. In A, a modified Miami plot shows features with positive beta values above the zero line and those with negative beta values below the zero line. The dark blue points indicate features with FDR q-value < 0.05 for data obtained for each column (C18 and HILIC). In B, the overlap in features associated with clinical AD and biomarker positive status at nominal p < 0.05 (light blue and dark blue points) for each column. In C, the metabolic pathways, with Fisher’s exact test p < 0.3, enriched by features nominally associated with the clinical AD and biomarker positive status. An asterisk indicates pathway that were significantly enriched (p < 0.05).

We identified 107 metabolites nominally associated (p<0.05) with both clinical AD and biomarker positive status for AD. Metabolites associated with being biomarker positive for AD were enriched in tyrosine and urea cycle/amino acid metabolism pathways. Dodecanoylcarnitine (adj p=0.009) and Ramipril (adj p=0.02) were the top analytes associated with clinical AD while LysoPC(18:0) (adj p=0.02) and creatinine (adj p=0.02) were associated with biological AD. 151 metabolites with a level 1-3 confidence score for annotation were associated (adj p<0.05) with at least one measured plasma biomarker (Figure 2, Supplementary Table 1). The strongest association observed was 3-oxododecanoic acid with αβ42/αβ40 ratio (adj p= 4.03E-23). Oxalic acid and hexacosanoyl carnitine were also strongly associated with αβ42/αβ40 ratio. Metabolites associated with αβ42/αβ40 ratio were enriched in heparan sulfate, chondroitin sulfate and keratan sulphate degradation processes. Increased sulfation and heparan sulfate proteoglycan degradation have been widely reported in AD related neuritic plaques previously.

**Figure 2.**
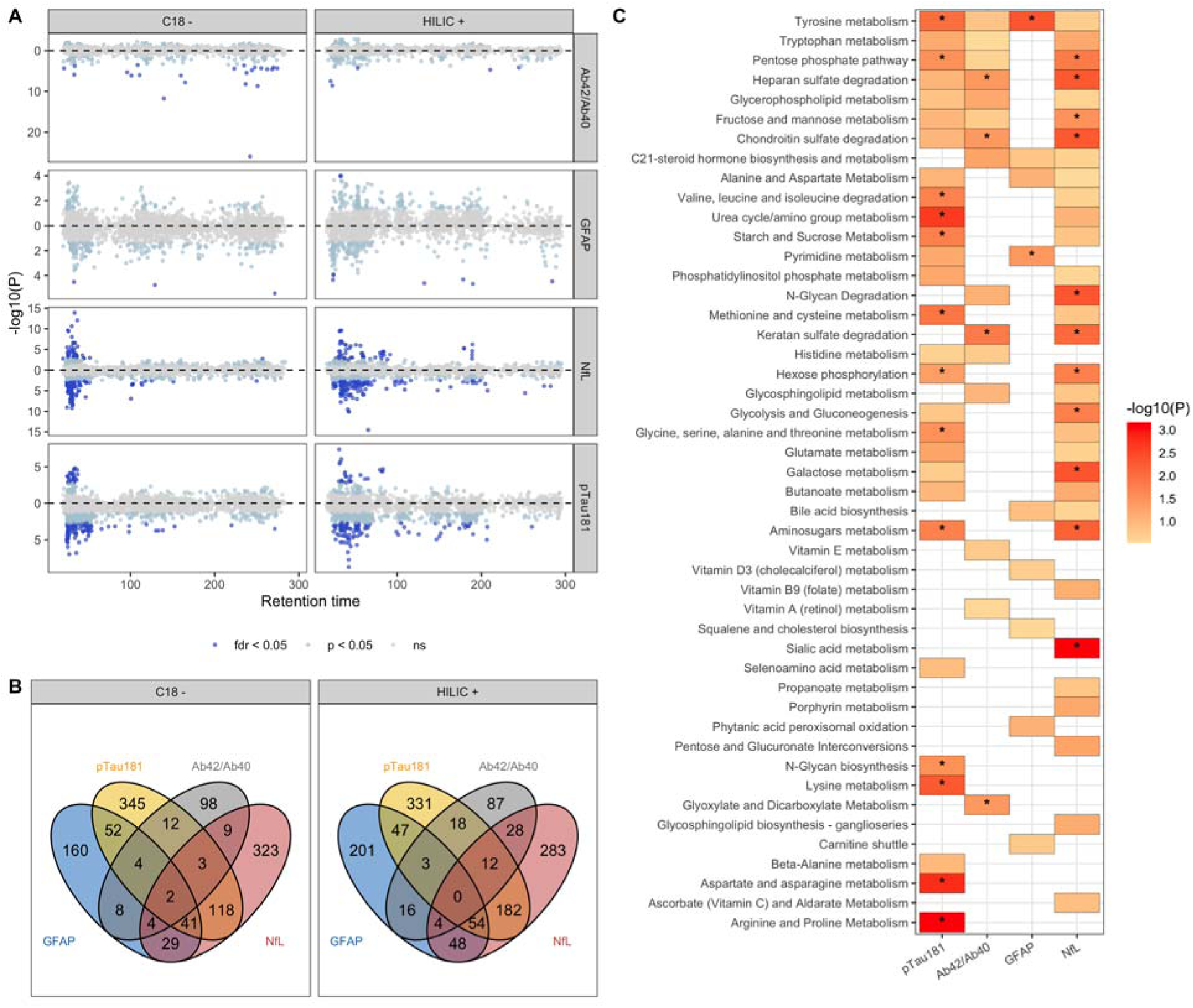
Metabolic features and pathways associated with plasma-based biomarkers of AD. In A, a modified Miami plot shows features with positive beta values above the zero line and those with negative beta values below the zero line. The dark blue points indicate features with FDR q-value < 0.05 for data obtained for each column (C18 and HILIC). In B, the overlap in features associated with the four biomarkers at nominal p < 0.05 (light blue and dark blue points) for each column. In C, the metabolic pathways, with Fisher’s exact test p < 0.3, enriched by features nominally associated with the biomarkers. An asterisk indicates pathways that were significantly enriched (p < 0.05). αβ42/ αβ40: ratio of A _42_ to A _40_ measured in plasma, GFAP: glial fibrillary acidic protein, NfL: neurofilament light chain, pTau181: tau phosphorylated at threonine-181.

Valyl serine, creatinine and citrulline were among the 76 well annotated metabolites associated with plasma levels of P-tau181. Several lysophosphatidylcholines (lysoPCs) including LysoPC(22:6), LysoPC(18:0) and LysoPC(20:4) were inversely associated after multiple testing correction with plasma P-tau181 levels (Supplementary Table 1). Metabolites associated with P-tau181 levels were enriched in several essential amino acid metabolism pathways including tyrosine, arginine and proline metabolism, valine, leucine, and isoleucine degradation, and aminosugars, starch and sucrose metabolism. Lysine metabolism, aspartate and asparagine metabolism and arginine and proline metabolism were enriched only among P-tau181 associated metabolites.

406 metabolites were associated with NfL levels in plasma, of which 107 were annotated with Level 1-3 confidence (Figure 2, Supplementary Table 1). Gluconate (adj p= 4.56E-11) and Arabinose (adj p= 2.93E-08) in the pentose pathway and DL-2-Aminoadipic Acid (adj p=1.53e-09) in Lysine/Glutamate metabolism were the top metabolites associated with NfL. Metabolites associated with NfL shared common pathways with those associated with αβ42/αβ40 ratio including heparan sulphate, chondroitin sulphate and keratan sulphate degradation. Pentose phosphate, aminosugars metabolism and hexose phosphorylation pathways were shared by metabolites associated with both NfL and P-tau181. Sialic acid metabolism was observed only amongst NfL associated metabolites.

Increased creatinine levels were associated with biological AD, increased amounts of plasma P-Tau181 and NfL. Several LysoPCs were observed to decrease in biological AD, and with increased amounts of plasma NfL and P-tau181 levels. Only LysoPC(18:1) was found to increase with increased levels of GFAP, although it was observed to decrease with increased P-tau181 and NfL levels indicating a time dependent abundance depending on the disease stage. **Metabolites in patients with discordant biological and clinical status of AD.** First we compared healthy participants with no negative biomarker diagnosis (BM-/Control) with the other three groups (BM+/Case, BM+/Control and BM-/Case) (Figure 3, Supplementary Figure S8). LysoPCs, in particular, LysoPC 22:6 (DHA) and LysoPC 20:5 (EPA) are reduced in clinical AD and biomarker positive patients. They are depleted the most in patients with both biological and clinical diagnosis of AD. Similarly, creatinine is increased in BM+ and clinical AD patients and is highest in patients with elevated Ptau-181. Tyrosine metabolism is enriched amongst metabolites that are elevated in patients with either clinical or biological AD. Glycosphingolipid metabolism is altered only in patients with only clinical diagnosis of AD and urea cycle and amino group metabolism is altered in biological AD patients.

**Figure 3.**
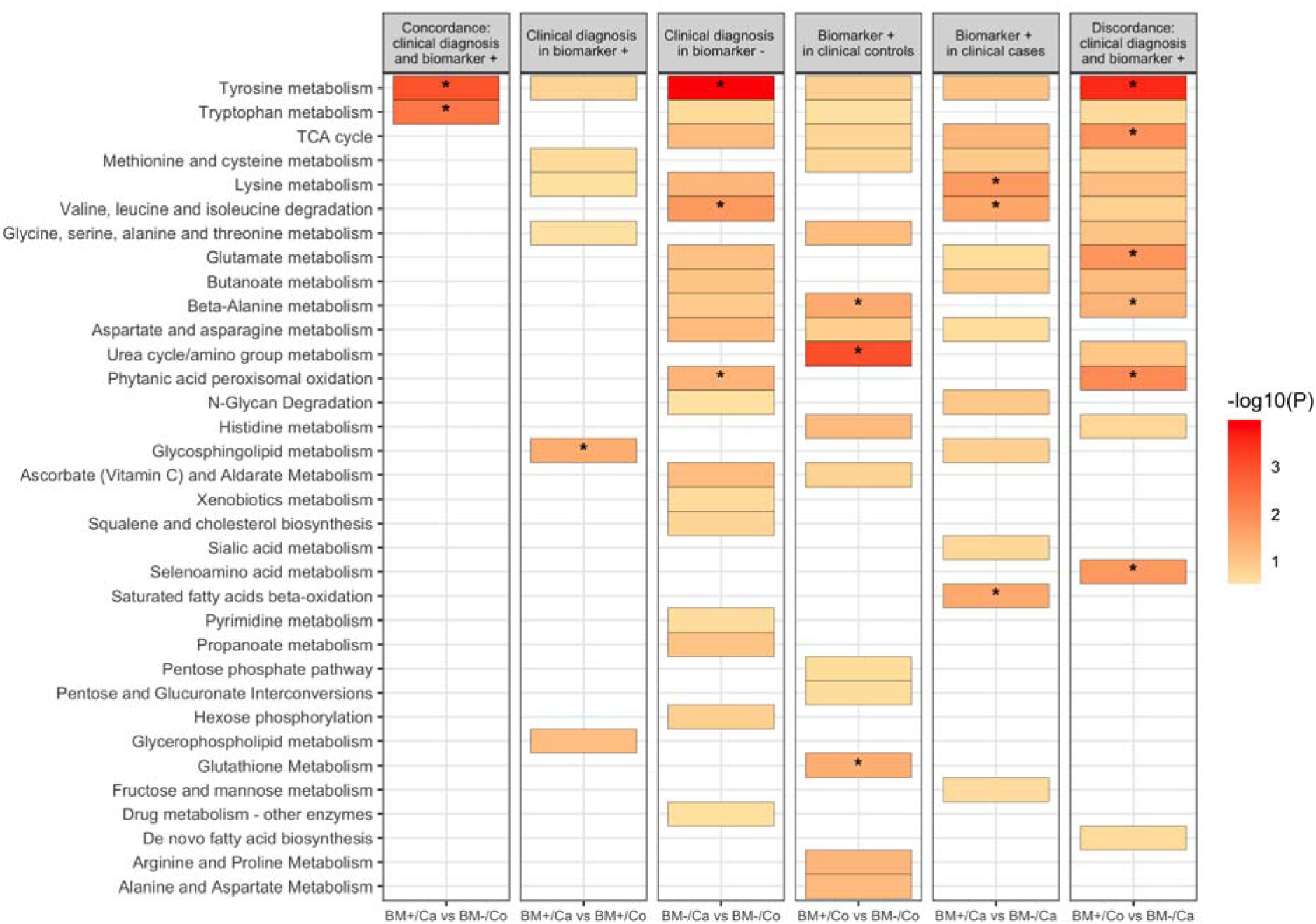
Subgroup analysis based on clinical diagnosis and biomarker positive status. Results from multinomial and regular logistic regression were used to determine metabolic pathways enriched. A colored box indicates an enriched pathway with Fisher’s exact test p-value < 0.3 while an asterisk indicates statistically significant enrichment (p < 0.05).

### Co-abundance analysis of metabolites

We clustered co-abundant metabolites using WGCNA independently on metabolites detected in the HILIC and C18 columns. WGCNA identified 18 and 15 co-abundant metabolic color-coded modules in HILIC and C18 columns respectively with at least 30 metabolites (Supplementary Figure S2). We then tested association of each module with clinical and biological AD and levels of the plasma biomarkers (Figure 4A). Purple module was negatively associated with biological AD (adj p=9e-05) while black module was associated with P-tau181 levels (adj p=3e-04). Salmon and greenyellow modules were associated both with biological AD and P-tau181 levels. Enrichment analysis of the metabolites co-abundant in the purple module found that fatty amides (adj p=5e-03), Glycerophosphocholines (adj p=5e-03) and sphingoid bases (adj p=5e-03) were over-represented in the module (Figure 4B). Glycerophosphocholines (adj p=3e-22) were also significantly enriched in the greenyellow module, while amino acids and peptides were the top group over-represented in the black (adj p=1.31e-16) and salmon modules (9.22E-07).

**Figure 4.**
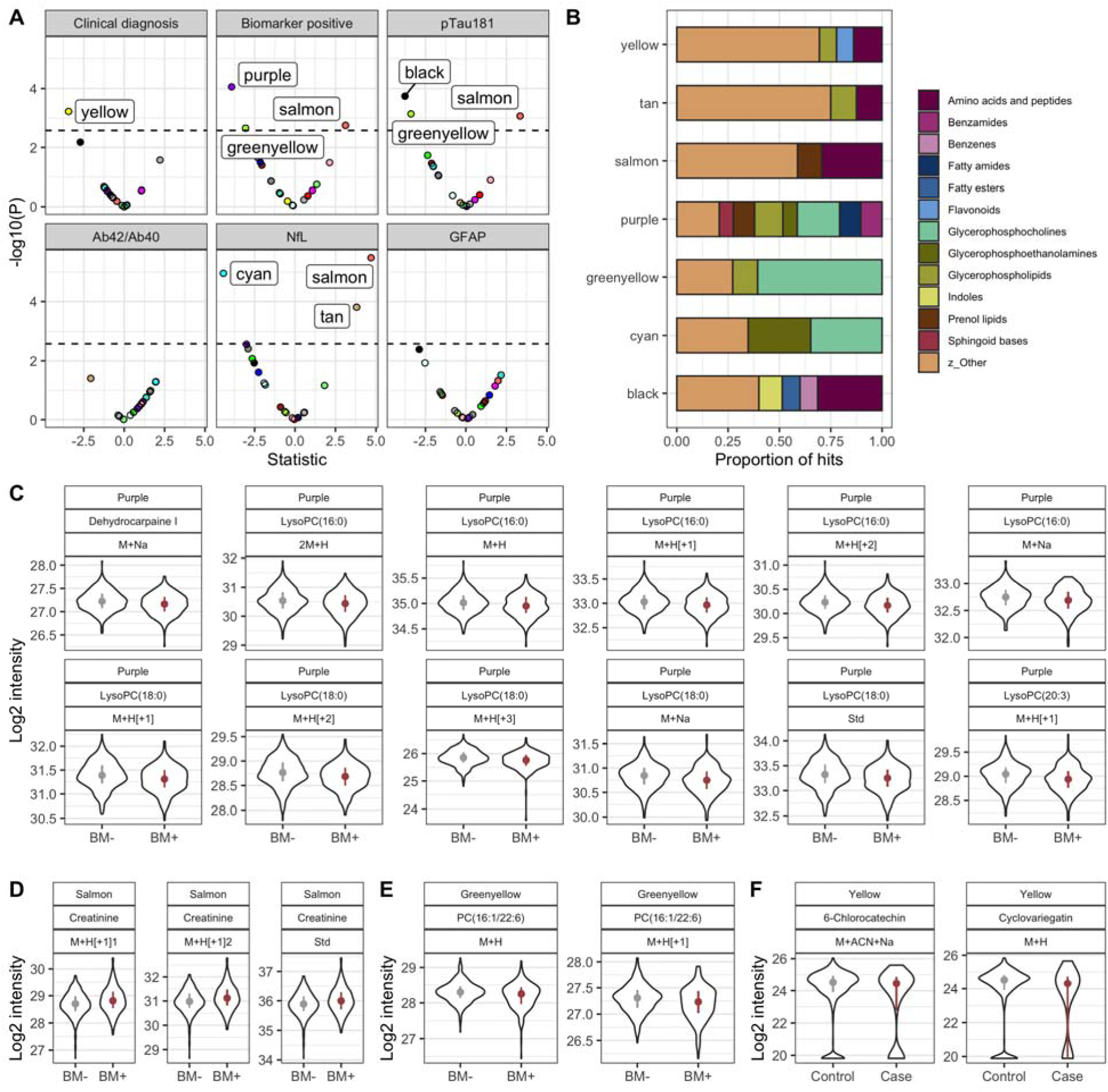
Results from co-expression analysis using data from the HILIC column. In A, the volcano plot shows metabolic modules significantly associated with clinical AD, biomarker positive status and AD biomarkers using Bonferroni adjusted p-value. In B, the chemical classes enriched by module member metabolic features present at a proportion of at least 6.5%. In C, module hub members of the purple module with KME > 0.6 and associated with biomarker positive status at FDR q-value < 0.05. In D, module hub members of the salmon module with KME > 0.6 and associated with biomarker positive status at FDR q-value < 0.05. In E, module hub members of the greenyellow module with KME > 0.6 and associated with biomarker positive status at FDR q-value < 0.05. In F, module hub members of the yellow module with KME > 0.6 and associated with clinical AD at FDR q-value < 0.05.

We then identified the hub metabolites that are most connected to other metabolites in the purple, salmon and greenyellow modules. Interestingly, 12 lysoPCs were hub metabolites in the purple module and all of them were more abundant in biomarker negative participants compared to individuals who were biomarker positive and defined to have biological AD (Figure 4C). Phosphatidylcholines (PC) and LysoPCs were also the hub metabolites in the greenyellow module and (Figure 4E, Supplementary table 2) and were also more abundant in healthy participants compared to biomarker positive patients. Creatinine was the most connected metabolite in the salmon module and as previously described, was increased in AD patients compared to controls.

### LysoPCs association with AD biomarkers

Both MWAS and WGCNA detected lysoPCs to be significantly associated with biological AD, P-tau181 and NFL levels. Thus, we tested joint association of all lysoPCs with clinical and biological AD by constructing lysoPC principal components. We constructed PCs for the 55 lysoPCs detected by HILIC and C18 columns found that the first five PCs explained over 75% of the variance (Supplementary Figure S3). We tested association of the first five PCs together in a regression model adjust for age and sex (Figure 5A). PC1 and PC4 were associated with biological AD whereas PC5 was associated with clinical diagnosis of AD. PC1 and PC5 were decreased in AD patients whereas PC4 was increased in disease. Further stratifying participants by presence of absence of *APOE* ε*4* allele (Figure 5B), we found that PC1 and PC5 were protective of biological AD and clinical AD respectively, only in *APOE* ε*4* non-carriers, whereas risk conferred by PC4 was restricted to *APOE* ε*4* carriers. We investigated the loadings of the LysoPCs on PCs 1,4 and 5 and particularly focused on LysoPCs that have poly unsaturated fatty acids (PUFAs) at the sn-1 and sn-2 positions (Figure 5C, Supplementary Figure S6). LysoPCs that carry eicosapentaenoic acid (EPA), docosahexaenoic acid (DHA) and arachidonic acid (AHA) are positively correlated with PC1 and hence decreased in biological AD, particularly in *APOE* ε*4* non-carriers. Both DHA and AHA were negatively correlated but EPA was positively correlated.to PC4 which increased risk of biological AD. We also tested the correlation between PCs 1,4 and 5 with circulating PUFAs in plasma (Supplementary Figure S5). PC1 was positively correlated with circulating levels of EPA and AHA, PC4 was negatively correlated with most measured plasma PUFAs and PC5 was positively correlated with linolenic acid, EPA and DHA and negatively correlated with AHA. This indicates that LysoPCs might play a role in AD biology in conjunction with *APOE*.

**Figure 5.**
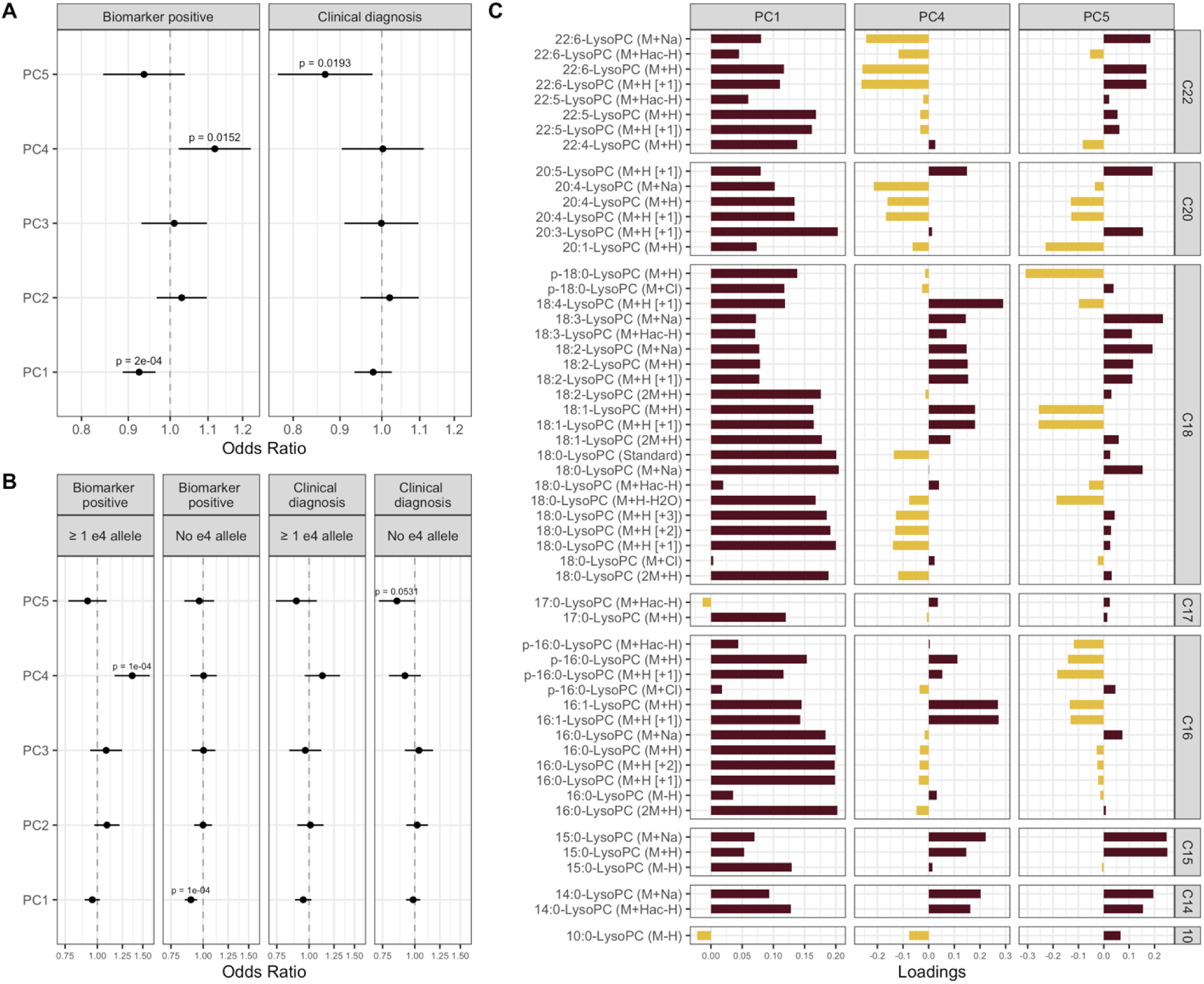
Lysophosphatidylcholines (LysoPCs) associated with clinical AD and biomarker positive status. In A, the odds ratio (point) and confidence interval (whiskers) of PC1 – 5 in relation to biomarkers positive status and clinical AD. In B, the results from analysis stratified by *APOE-4* allele status. In C, the loadings of LysoPCs on the three PCs (PC1, PC4, and PC5) significantly associated with biomarkers positive status or clinical AD.

### LysoPC analysis in the ROSMAP cohort

To determine the generalizability of our results we examined the association between lysoPCs and AD pathology in the ROSMAP cohort. We used the metabolomics data derived from 110 brain samples in the ROSMAP cohort to test association of LysoPCs and phosphatidylcholines (PCs) with pathological definition of AD, amyloid burden, tangle density, and global pathology. First, we detected 14 LysoPCs and 13 PCs in the ROSMAP cohort. We constructed principal components from the LysoPCs and tested association with AD pathology (Supplementary Figure S7). We identified that PC3 is positively associated with increased tangle density, global pathology and a pathological diagnosis of AD. Of the LysoPCs carrying PUFAs, we only detected AHA in the ROSMAP cohort. AHA is increased in tangles, global pathology and pathological definition of AD. Three phosphatidylcholines were negatively associated with amyloid burden and tau tangles implying that LysoPCs and PCs are reduced in post-mortem AD brains. These findings are consistent with plasma LysoPCs observation in the EFIGA cohort.

## Discussion

We investigated the association of metabolites with plasma biomarkers and clinical diagnoses in a cohort of Caribbean Hispanics to identify metabolic pathways associated with hallmarks of AD pathology. Two of the most notable findings were that metabolite profiles differed when a clinical diagnosis was used versus a validated plasma biomarker-based diagnosis and that lysoPCs, which have been reported in recent studies in AD^59,60^, were identified in our unbiased approach in a Hispanic population.

LysoPCs were associated with both quantitative levels of plasma P-tau181 and biological AD (defined by P-tau181). Co-abundance analysis revealed P-tau181 association of metabolic modules that harbor several LysoPCs as hub metabolites, suggesting a critical role in disease pathogenesis. Several studies have observed lower levels of lysoPCs in the brains, CSF and plasma of AD patients^61–72^. These changes often involve lysoPC species, particularly those that bind to anti-inflammatory PUFAs being decreased in patients with AD. Some lysoPC species have been implicated in promoting neurotoxicity and inflammation^73–75^. They can induce oxidative stress, impair mitochondrial function, and activate immune cells, leading to neuronal damage and death. LysoPCs are also involved in dysregulation of lipid metabolism these disturbances. The breakdown of phosphatidylcholine, a major lipid component of cell membranes, can generate lysoPCs. Disruptions in enzymes involved in this process, such as phospholipase A2 (PLA2), have been observed in AD and may contribute to altered lysoPC levels^76–78^.

We observed a differential effect of LysoPCs within *APOE* ε4 carriers and non-carriers. The risk conferred by LysoPCs was restricted to *APOE* ε4 carriers, while the protective effects were significant within *APOE* ε4 non-carriers. We previously showed significant differences in metabolic profiles in a small multi-ethnic AD cohort and these differences remained when the analysis was restricted to *APOE* ε4 carriers^79^. *APOE* ε4 carriers tend to exhibit higher levels of specific LysoPC species in CSF, plasma, and brain tissue compared to non-carriers^80–83^. Elevated levels of certain LysoPCs in *APOE* ε*4* carriers have been linked to increased Aβ deposition, tau phosphorylation, and neuroinflammation. Distinct patterns of LysoPC alterations have been observed in *APOE* ε4 carriers compared to non-carriers.

We also found essential amino acids metabolism (tryptophan and tyrosine) were associated with clinical and biological diagnosis of AD. Urea cycle/amino group metabolism was associated with only the biological diagnosis of AD. Tyrosine is an essential amino acid and plays a crucial role in the synthesis of catecholamines. Limited research has focused on measuring tyrosine levels specifically in the AD patient brains but administering tyrosine orally can enhance memory and cognitive function^84^. Tryptophan is an essential amino acid and a precursor for the synthesis of serotonin, a neurotransmitter involved in mood regulation and cognition. Alterations in tryptophan metabolism may impact serotonin availability in the brain and contribute to AD pathophysiology, particularly (Aβ) pathology. Aβ accumulation can disrupt tryptophan metabolism, leading to altered levels of tryptophan and its metabolites. Conversely, tryptophan metabolites, such as kynurenic acid, can affect Aβ aggregation and clearance, potentially influencing disease progression. Interestingly Tryptophan levels in plasma were associated with clinical diagnosis of AD and were also mildly correlated with CSF levels (correlation=0.24, Table 2).

Heparan sulfate, chondroitin sulfate and keratan sulphate degradation processes were associated with Aβ42/40 ratio. Heparan sulfate, chondroitin sulfate, and keratan sulfate are types of glycosaminoglycans (GAGs) or sulfated carbohydrates that are found in the extracellular matrix of cells. GAGs have been reported in accumulation and clearance of in Aβ. Heparan sulfate proteoglycans (HSPGs) are a type of protein with heparan sulfate chains that interact with Aβ and can contribute to the formation of amyloid plaques. Chondroitin sulfate proteoglycans (CSPGs) and HSPGs have been implicated in the regulation of Aβ clearance. These sulfated glycosaminoglycans can interact with various proteins involved in the clearance of Aβ, including neprilysin and insulin-degrading enzyme. Disruption of the balance between Aβ production and clearance, partly mediated by GAGs, may contribute to the accumulation of Aβ in AD. GAGs can interact with various inflammatory molecules, including cytokines and chemokines, and modulate neuroinflammatory processes in AD. Chondroitin sulfate and heparan sulfate chains present on proteoglycans can act as binding sites for inflammatory molecules, contributing to the activation of immune cells and the generation of a pro-inflammatory environment in the brain.

Taken together these results suggest that understanding metabolic heterogeneity in AD pathogenesis may enable identification of biological mechanisms for specific subgroups with the disease and that it is essential to combine biochemical analysis with biomarkers of disease. Specifically, identification of metabolic pathways associated with plasma biomarkers might indicate biological mechanisms underlying AD pathology at different stages of the disease. We observed common metabolic pathways perturbed in clinical AD and elevated Aβ42/40 ratio, indicating that these metabolites might be involved in both amyloidogenic and later (clinical) stages of the disease. Similarly, distinct set of metabolites were observed in association with elevated P-tau181 and NfL levels, suggesting processes that might be involved both in neurofibrillary change and neurodegeneration. However, more investigation specifically with longitudinal measures of biomarkers and metabolic assessments are needed to disentangle the metabolic cascades in different stages of disease progression. Finally, this study demonstrates the ability of high-resolution mass spectrometry-based untargeted metabolomics to reveal biochemical differences in participants with differential plasma biomarker profiles and to identify metabolic perturbations in different stages of the disease. This has the potential to open up a new era of biochemically-based discovery in AD.

## Data Availability

All data produced in the present study are available upon reasonable request to the authors

## Acknowledgements

EFIGA study is supported by NIA grants R56AG063908, R01AG067501 and RF1AG015473. We acknowledge the services of CEDIMAT for collaborating with sample collection and processing in the EFIGA cohort.

The metabolomics core that generated the metabolomics data for the project is supported by the National Center for Advancing Translational Sciences grant-5UL1TR001873.

