## Supplementary Figures for "Lysophosphatidylcholines are associated with P-tau181 levels in early stages of Alzheimer’s Disease"

**Supplementary Figures and Tables**

Fig S1: PCA plot showing PC1 vs PC2 of all metabolites that pass QC.

29 individuals identified as outliers were removed from the analysis.

HILIC +

C18 -

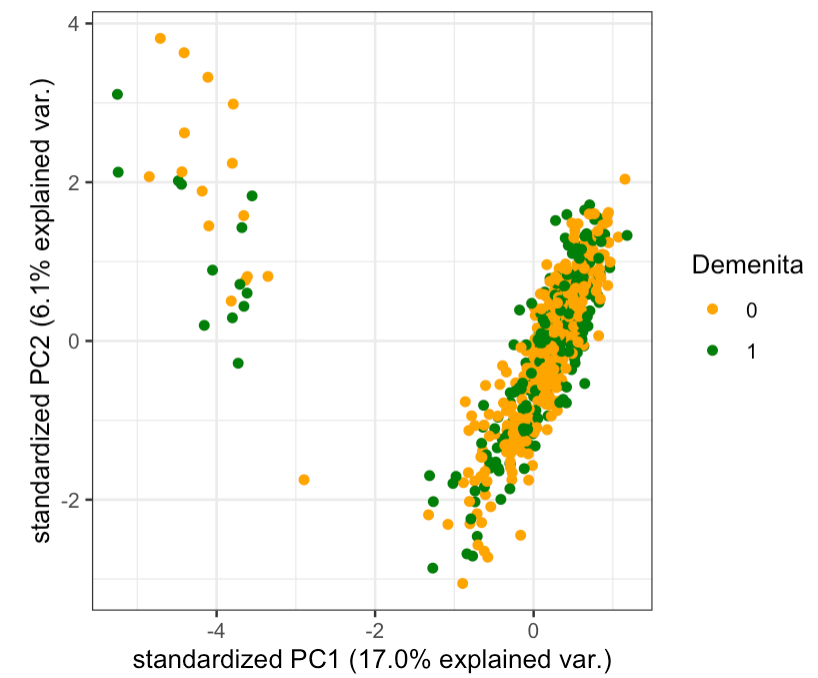

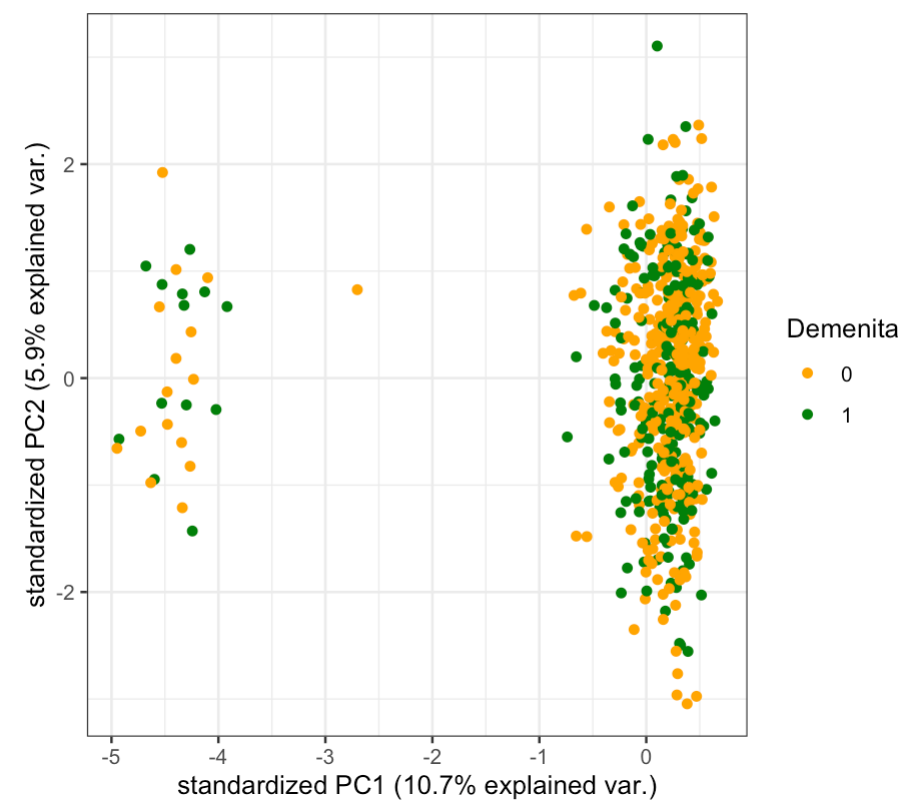

Fig S2: Modules of metabolites detected by WGCNA

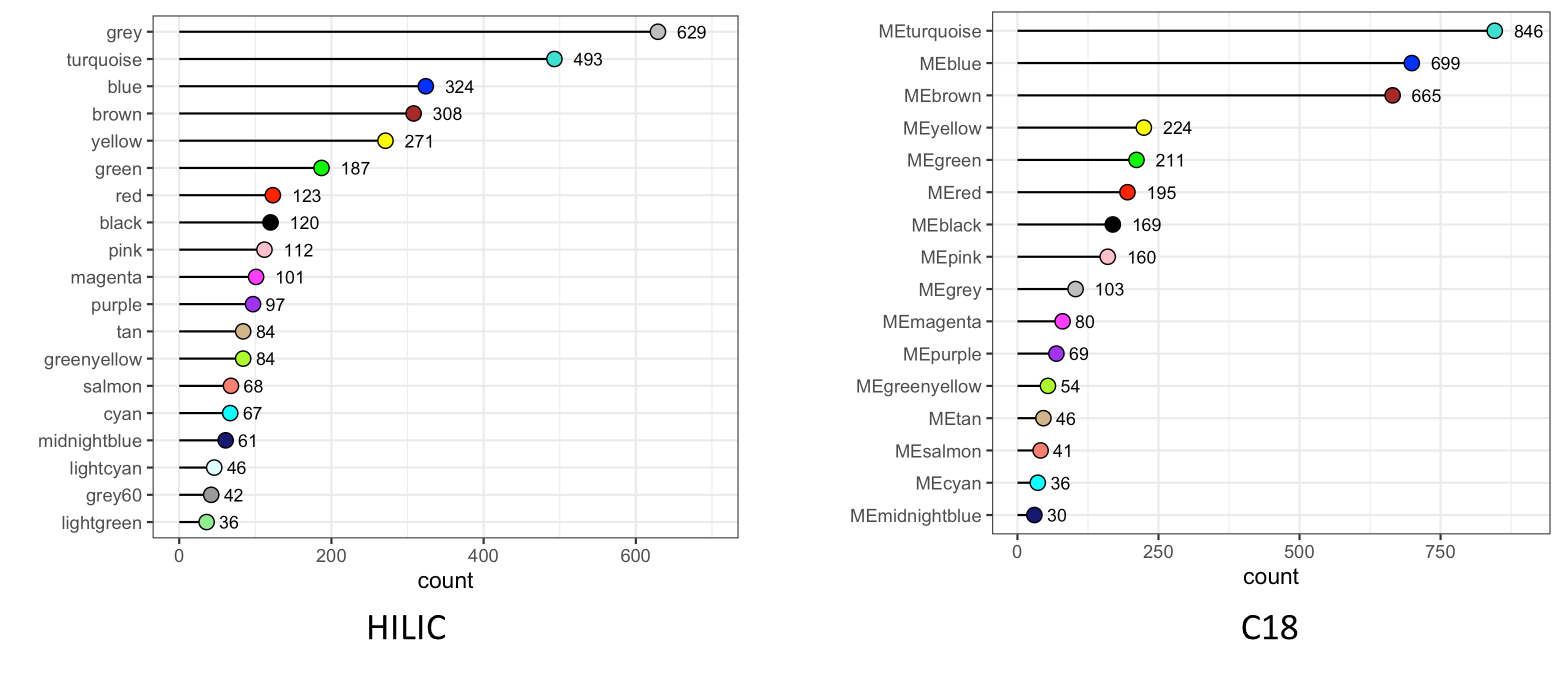
grey module contains metabolites not clustered with any other metabolites.

Fig S3. Scree plot of LysoPC components

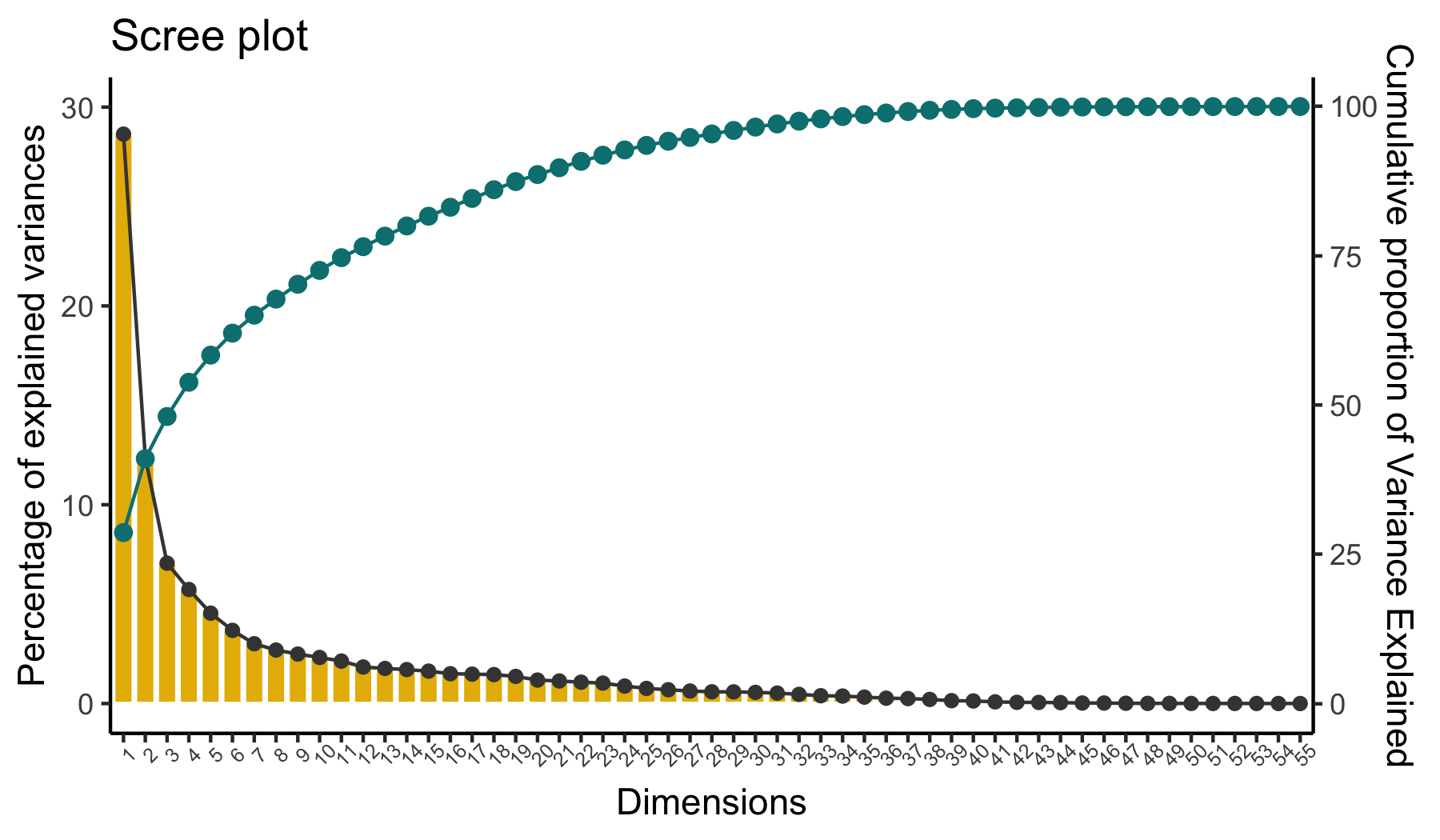

**Figure S4. Results from co-expression analysis using data from the C18 column.** In A, the volcano plot shows metabolic modules significantly associated with clinical AD, biomarker positive status and AD biomarkers using Bonferroni adjusted p-value. In B, the chemical classes enriched by module member metabolic features present at a proportion of at least 6.5%. In C, module hub members of the cyan module with KME > 0.6 and associated with biomarker positive status at FDR q-value < 0.1. In D, module hub members of the cyan module with KME > 0.2 and associated with clinical AD at FDR q-value < 0.05. In E, module hub members of the salmon module with KME > 0.6 and associated with clinical AD at FDR q-value < 0.05. In F, module hub members of the brown module with KME > 0.6 and associated with clinical AD at FDR q-value < 0.05. In G, module hub members of the greenyellow module with KME > 0.6 and associated with biomarker positive status at FDR q-value < 0.1.

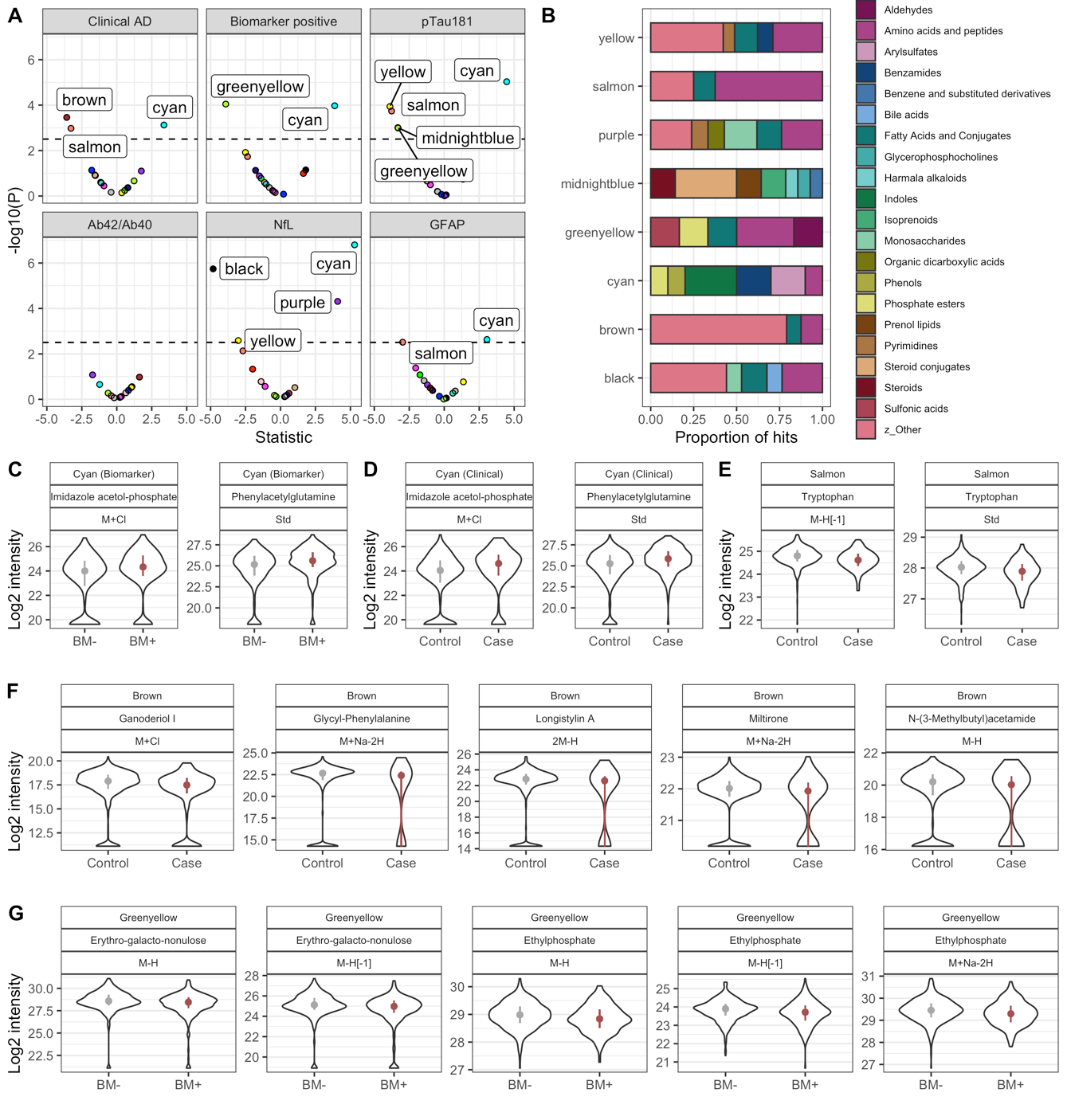

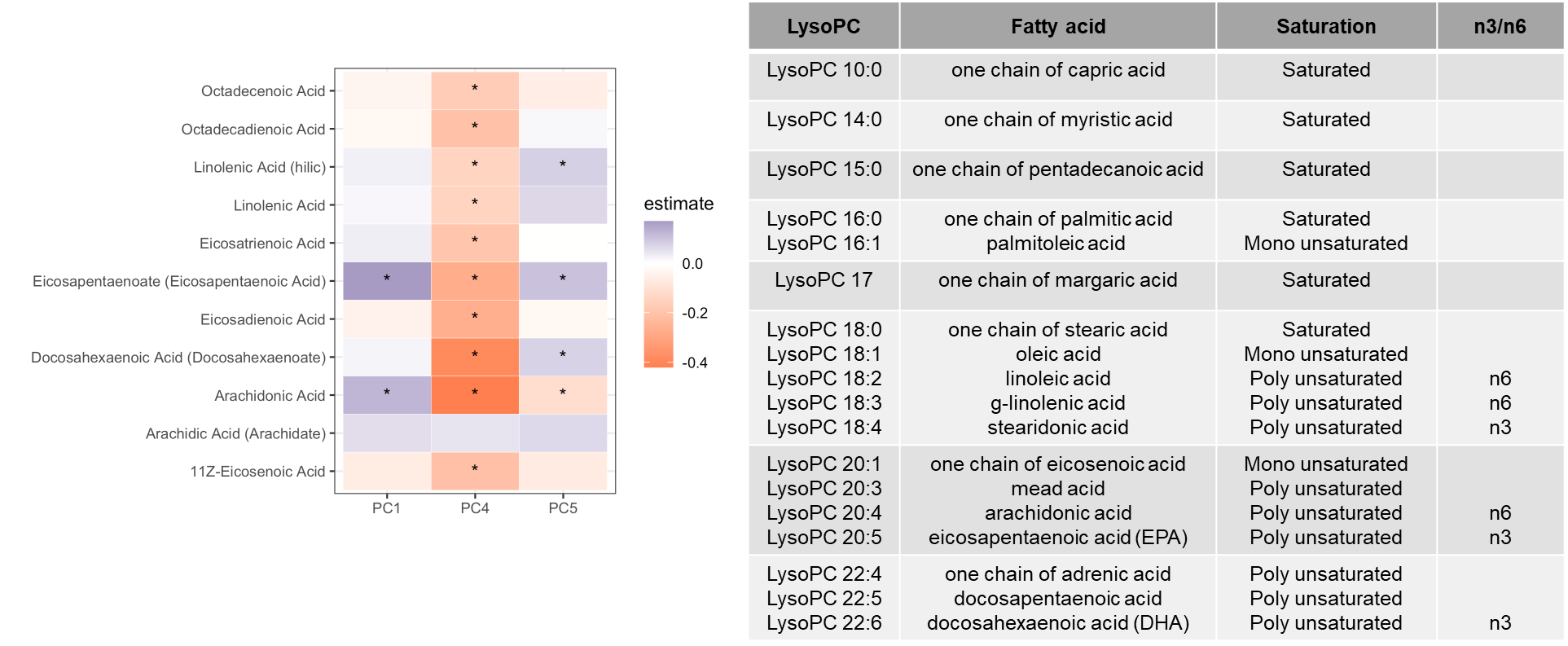
Supplementary Figure S5: Correlation of LysoPCs with circulating PUFAs in plasma

Supplementary Figure S7: Brain LysoPC association in ROSMAP cohort

S7-A First five principal components explain over 95% of the variance

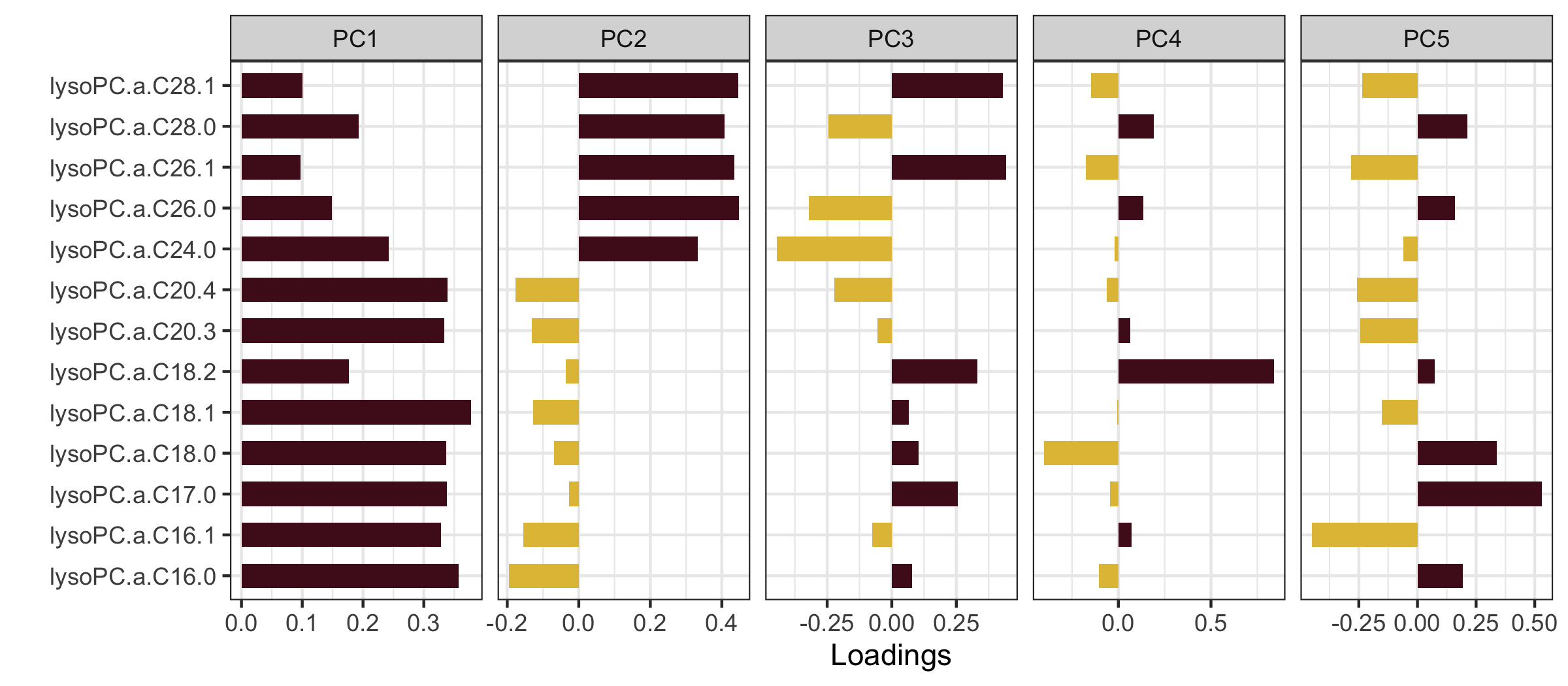

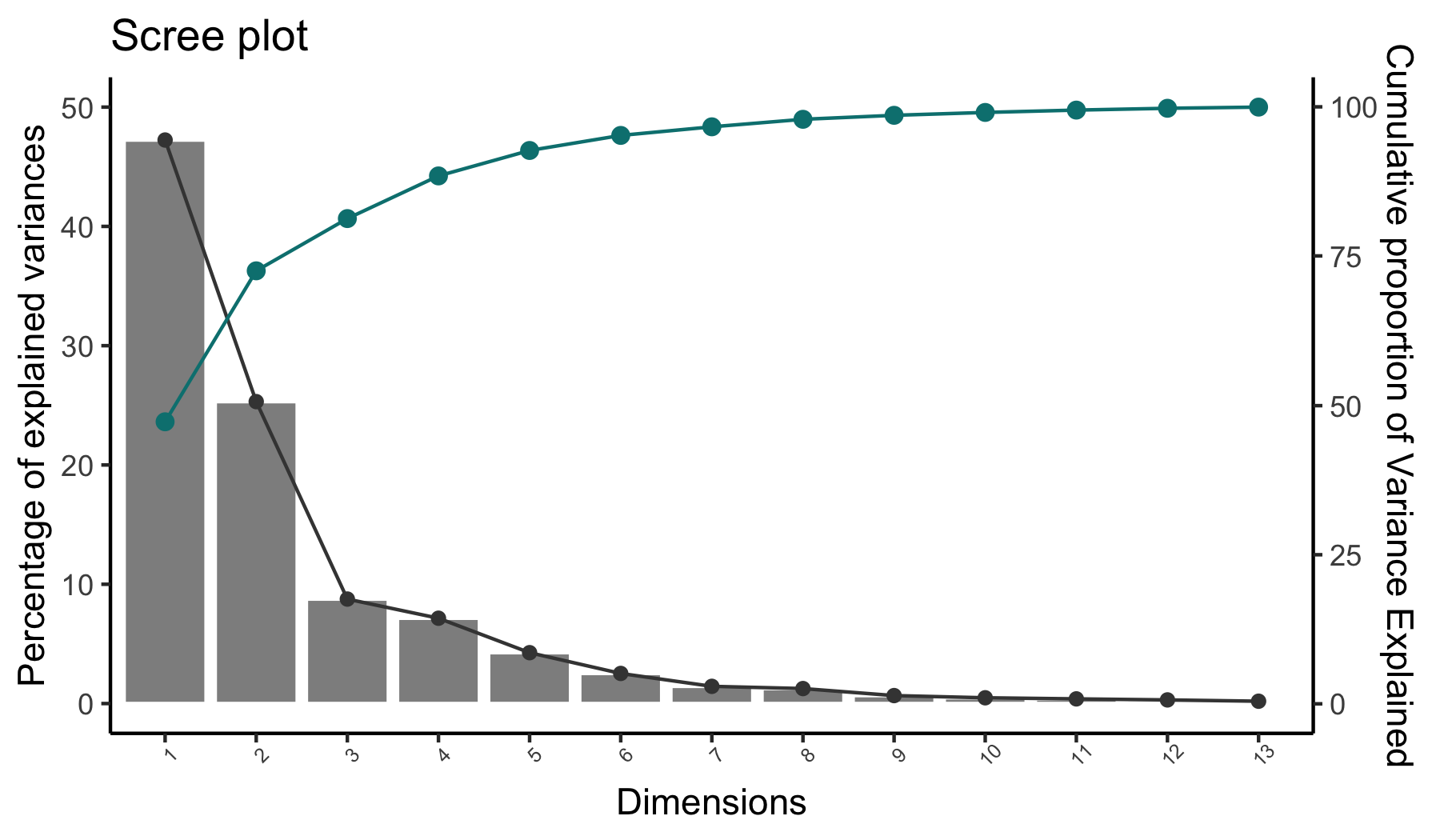

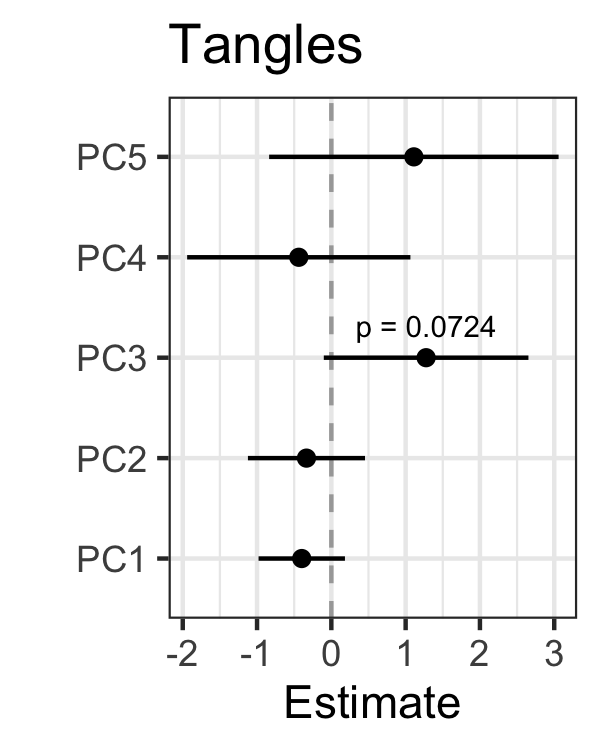

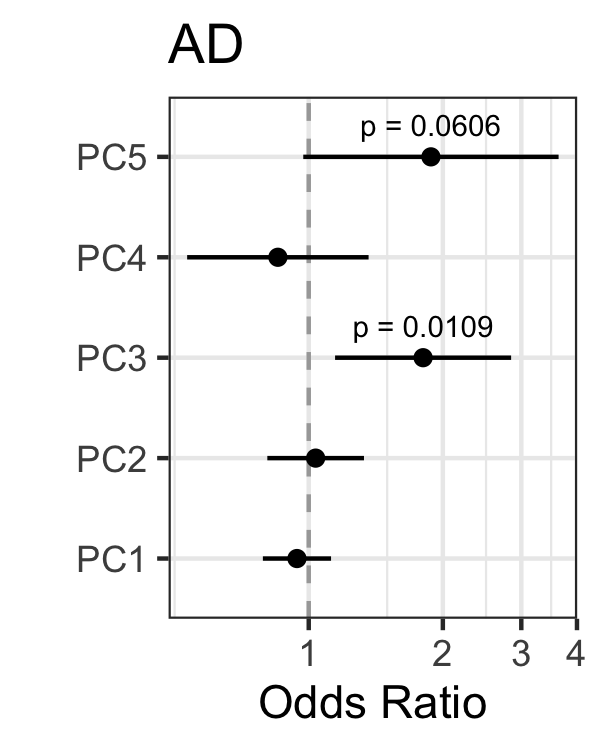

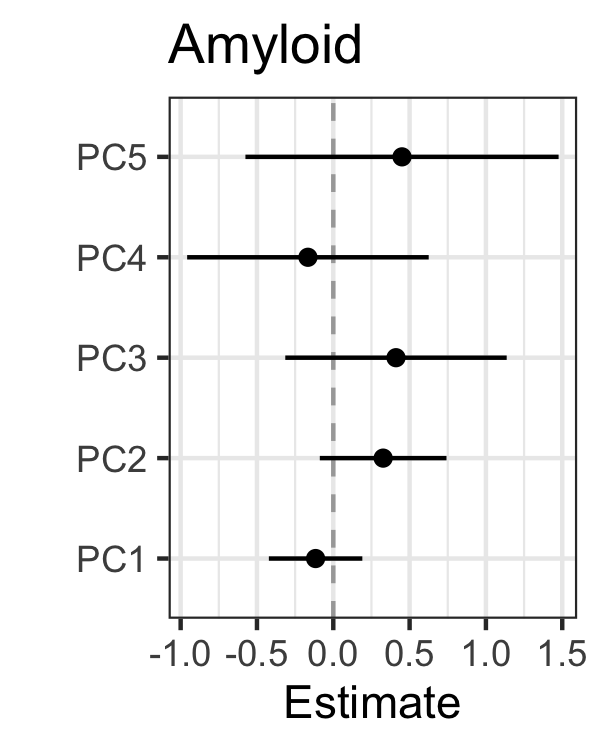

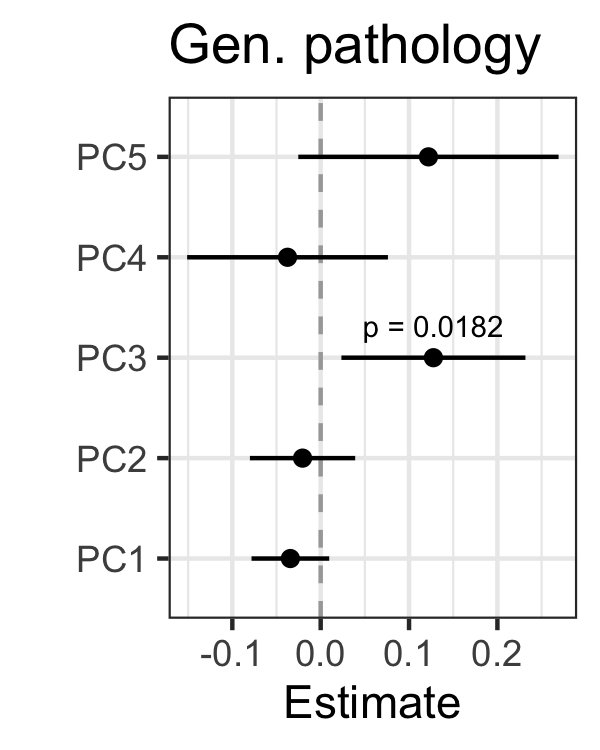

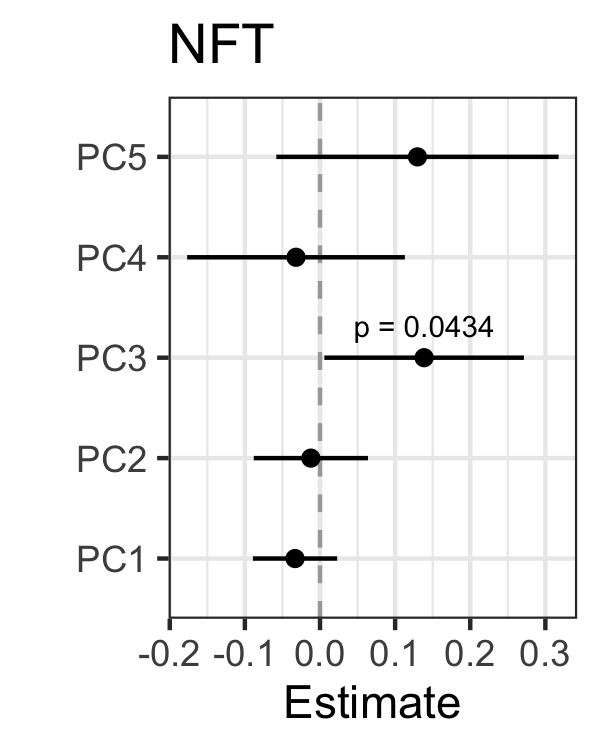

A

B

C

S7-B PC3 is nominally associated with pathological definition of AD, Tau Tangles, General pathology and Neurofibrillary tangles.

S7-C LysoPCs detected in the ROSMAP cohort, EPA and DHA carrying LysoPCs were not detected. **LysoPC.a.20.4 carries AHA**

Supplementary Figure S8: Levels of metabolites by Biomarker and clinical AD status

Top left= Creatinine, Top right= phenylacetylglutamine, Bottom left=LysoPC (20:5-EPA), Bottom Right=LysoPC (22:6-DHA)

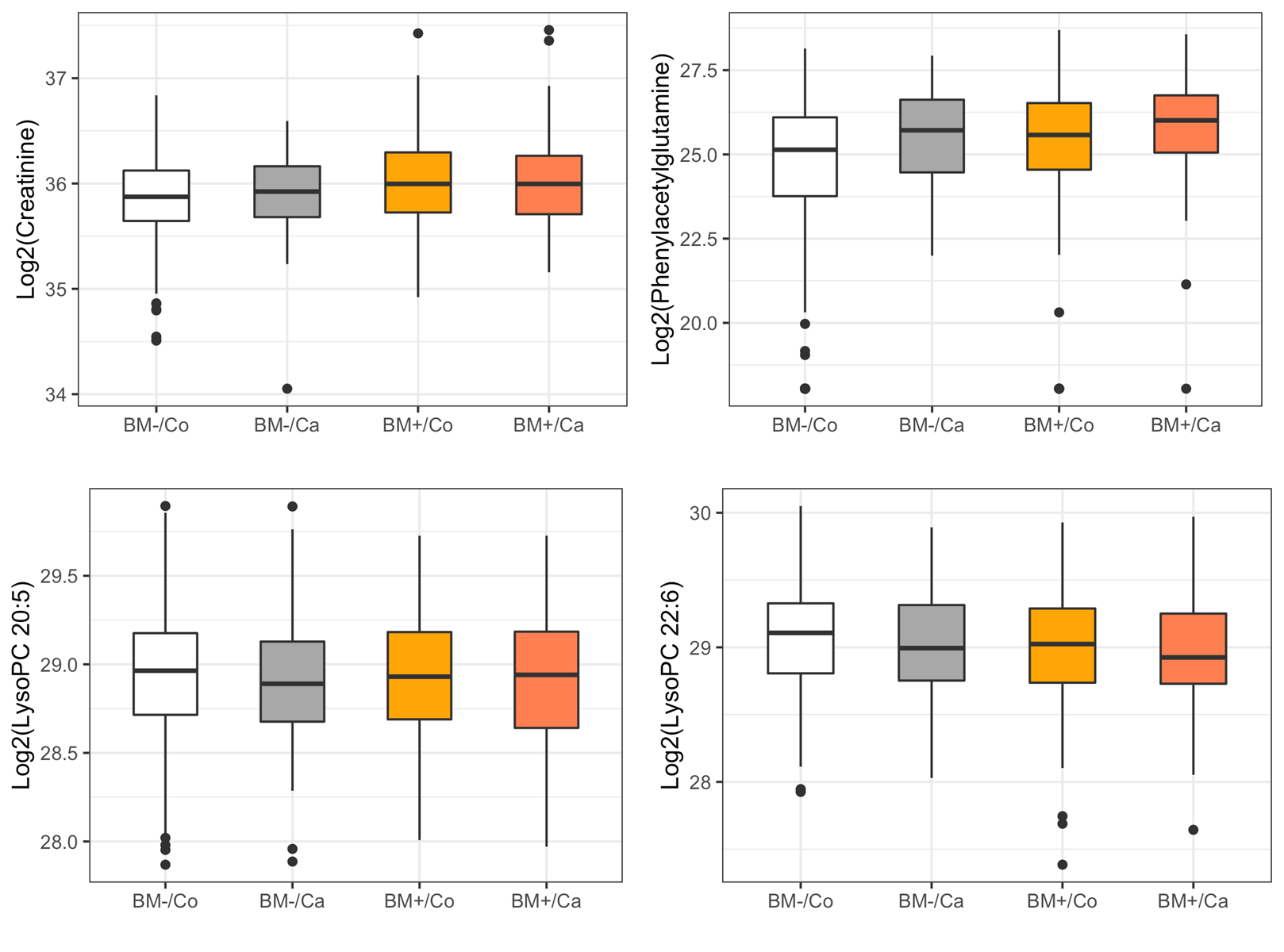
